# LiMAP-curvature: a simple model-free approach for analysing dose-finding studies

**DOI:** 10.1101/2022.10.30.22281725

**Authors:** Linxi Han, Qiqi Deng, Zhangyi He, Feng Yu

## Abstract

MCP-Mod (Multiple Comparison Procedure-Modelling) is an efficient statistical method for the analysis of Phase II dose-finding trials, although it requires specialised expertise to pre-specify plausible candidate models along with model parameters. This can be problematic given limited knowledge of the agent/compound being studied, and misspecification of candidate models and model parameters can severely degrade its performance. To circumvent this challenge, in the work, we introduce LiMAP-curvature, a Bayesian model-free approach for the detection of the dose-response signal in Phase II dose-finding trials. LiMAP-curvature is built upon a Bayesian hierarchical framework incorporating information about the total curvature of the dose-response curve. Through extensive simulations, we show that LiMAP-curvature has comparable performance to MCP-Mod if the true underlying dose-response model is included in the candidate model set of MCP-Mod. Otherwise, LiMAP-curvature can achieve performance superior to that of MCP-Mod, especially when the true dose-response model drastically deviates from candidate models in MCP-Mod.

## 1. Introduction

Characterising the dose-response relationship and finding the right dose are important but challenging in the pharmaceutical drug development process. Nearly half of failures in Phase III trials result in part from a lack of understanding of the dose-response relationship in Phase II trials (Sacks et al., 2014). Over the last decade, the Multiple Comparison Procedure-Modelling (MCP-Mod) method, developed by Bretz et al. (2005), has been increasingly popular for Phase II trials as it can provide superior statistical evidence for dose selection.

MCP-Mod is a two-step approach that combines MCP principles and modelling techniques. In the first MCP step, it establishes a dose-response signal (Proof of Concept, PoC), and in the second Mod step, it estimates the dose-response curve and target doses of interest. MCP-Mod overcomes shortcomings of traditional approaches for dose-finding studies (see e.g. Ting, 2006, for an excellent introduction). MCP-Mod requires the pre-specification of plausible candidate models and model parameters to capture model uncertainty, which however is primarily based on the limited knowledge of the agent/compound being studied, if it is available at all (Chen & Liu, 2020). Misspecification of candidate models and model parameters in MCP-Mod may cause a loss in power and unreliable model selection (see Saha & Brannath, 2019, and references therein).

Motivated by the limitations of MCP-Mod, non-parametric methods have gained popularity for detecting dose-response trends and estimating dose-response relationships in Phase II trials. These methods offer flexibility and adaptability to capture complex patterns in the data without imposing strong assumptions on the underlying functional form of the dose-response curve. West & Harrison (2006) introduced the normal dynamic linear model (NDLM), which leverages the flexibility of dynamic linear models to estimate the dose-response curve. By accommodating time-varying coefficients, NDLM can capture the dynamic nature of the dose-response relationship, allowing for more accurate and interpretable estimates. Kirby et al. (2009) applied cubic smoothing splines with generalised cross-validation for the smoothing parameter to estimate the dose-response curve.

However, these approaches still face challenges in incorporating prior knowledge about the curvature or shape of the dose-response curve. Determining the optimal level of smoothness or choosing the appropriate model can be subjective and dependent on the specific data at hand. To address these limitations, in this work, we develop a model-free Bayesian approach, which is a novel Bayesian hierarchical framework incorporating the total (in the *L*^2^ sense) curvature of the dose-response curve as a prior parameter. Our approach avoids the requirement of a set of pre-specified candidate models. The responses at the given set of doses are estimated through maximum *a posteriori* (MAP), with which we construct a test statistic to establish PoC through simulations. We can then estimate the dose-response relationship with simple interpolation.

The remainder of this work is organised as follows. We describe in detail our MAP approach with a curvature prior, abbreviated LiMAP-curvature, in Section 2. In Section 3, we evaluate the operating characteristics of LiMAP-curvature through simulations and compare its performance to that of MCP-Mod and smoothing spline. We present concluding remarks and future directions in Section 4.

## 2. Methods

We consider a trial with a total of *M* + 1 distinct doses *x*_0_, *x*_1_, … , *x*_*M*_ , where *x*_0_ represents placebo. We let *N*_*i*_ be the number of patients in dose group *i* and *f*(*x*) be the true dose-response function at dose *x*. We assume that *f*(*x*) is defined on the interval [0, 1] to align with the typical range of doses in pharmaceutical drug development where doses are often scaled or normalised within this range for convenience and comparability. For *i* = 0, 1, … , *M* and *j* = 1, 2, … , *N*_*i*_, we let *Y*_*ij*_ be the response observed for patient *j* allocated to dose *x*_*i*_. We assume that

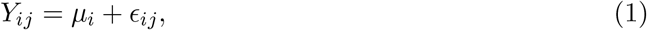

where *µ*_*i*_ = *f*(*x*_*i*_) denotes the mean response at dose *x*_*i*_, and 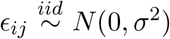 denotes the error term for patient *j* in dose group *i*. For the sake of simplicity, we assume that the variance *σ*^2^ is known and constant across all dose groups. This assumption is commonly used in different areas, including MCP-Mod (Bretz et al., 2005), BMCPMod (Fleischer et al., 2022) and Bayesian meta-analysis (Burke et al., 2018). See Fleischer et al. (2022) for further discussion on the assumption on the standard deviation *σ*.

In MCP-Mod, a set of plausible candidate models is required. This constrains the possible set of dose-response curves. However, with limited knowledge of the agent/compounds in Phase II trials, it is more likely to mis-specify the set of candidate models. The procedure for specifying candidate models is also somewhat cumbersome. So we would like to avoid the pre-specification of possible models beforehand, but still want to impose a certain degree of smoothness on the dose-response curve. To this end, we introduce the *L*^2^-total curvature

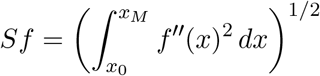

to measure how far the dose-response curve *f*(*x*) is from being a straight line. We will impose a half-normal *HN*(γ^2^) prior on *Sf* to give low prior probabilities to the dose-response relationship that is very curved, where the standard deviation *γ* controls the trade-off between the *L*^2^-total curvature of *f*(*x*) and fidelity to data ***Y*** . To capture an appropriate level of curvature and allow for optimal model performance, we assign a hyperprior for *γ* with a half-normal distribution *HN*(*τ* ^2^), which reflects our prior beliefs about the likely range of values for *γ* before observing any data. By incorporating a hyperprior, we introduce additional uncertainty and flexibility into the prior distribution of *Sf* , influencing the estimation of the dose-response curve. The specification of the standard deviation *τ* will be discussed in Section 4.

Given the dose-response function *f*(*x*) being available at *M* + 1 distinct doses, for *i* = 1, 2, … , *M* − 1, we have

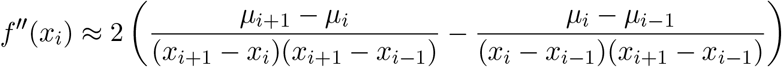

through the second-order central difference scheme (Burden et al., 2015), therefore the *L*^2^-total curvature *Sf* being approximated through numerical integration with

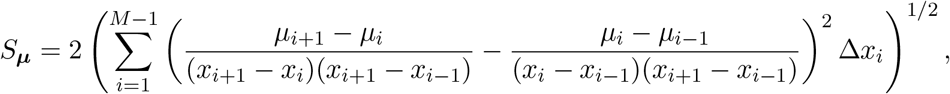

where Δ*x*_*i*_ = (*x*_*i*+1_ − *x*_*i*−1_)/2 for *i* = 2, 3, … , *M* − 2 with Δ*x*_1_ = (*x*_2_ + *x*_1_)/2 − *x*_0_ and Δ*x*_*M*−1_ = *x*_*M*_ − (*x*_*M*−1_ + *x*_*M*−2_)/2.

We let

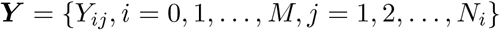

and define our Bayesian hierarchical model to be

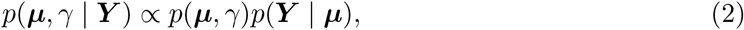

where the prior

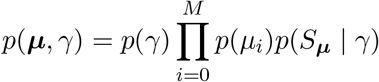

With

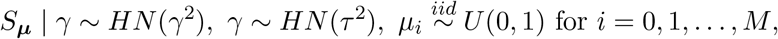

and the likelihood

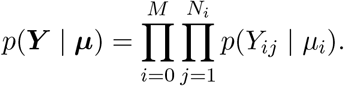

Suppose the standard deviation *τ* in the hyperprior for *γ* is pre-specified. The MAP estimates of all parameters in the model defined in Eq. (2) can be obtained by maximising the corresponding log-likelihood

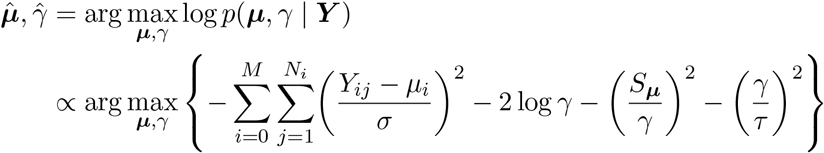

through a numerical optimisation algorithm like the Broyden–Fletcher–Goldfarb–Shanno (BFGS) method and its variants (see, e.g. Nocedal & Wright, 1999, for more details).

To establish PoC, we propose a test statistic

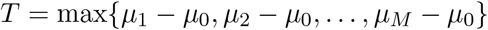

with hypotheses

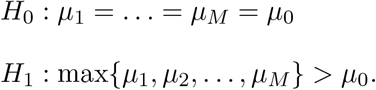

We define a significance level *α* for a dose-response signal, such that the corresponding critical value *c* satisfies

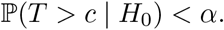

For a given *α*, we compute the critical value *c* via simulation

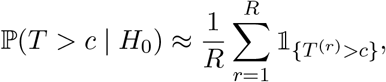

where *T* ^(*r*)^ is the test statistic computed from the data ***Y*** ^(*r*)^ simulated under the null hypothesis *H*_0_, *R* is the total number of replicates, and 𝟙_*A*_ is the indicator function equal to 1 if condition *A* holds and 0 otherwise.

Finally, the mean response estimates ***û*** can be linearly interpolated (or using a more sophisticated interpolation scheme) to obtain an estimate of the dose-response curve *f*(*x*), which will also yield an estimate of the target dose of interest. In this work, we only perform simple linear interpolation as we shall focus on the PoC stage of dose-finding trials.

## 3. Simulations

In this section, we assess the performance of LiMAP-curvature, in terms of power to detect dose-response signals as well as estimates of the dose-response curve and target doses of interest. We compare LiMAP-curvature to smoothing spline and MCP-Mod.

### 3.1 Simulation settings

We simulate randomised, double-blind, placebo-controlled, parallel-group trials with patients being equally allocated to placebo (0) or one of four active doses (0.15, 0.50, 0.80 and 1). We take the placebo effect to be 0 and the maximum treatment effect to be 0.5, respectively. We vary the sample size per dose group in {10, 20, 30, 40, 50, 60}. We choose one of 12 common dose-response shapes to be the true dose-response model. These models are plotted in Figure 1, with corresponding parameters summarised in Supplemental Material, Table S1. We simulate each patient’s response according to Eq. (1) with a standard deviation of *σ* = 1. For each of 72 combinations of parameters, consisting of sample size and dose-response shape, we run 10,000 simulated trials.

**Figure 1:**
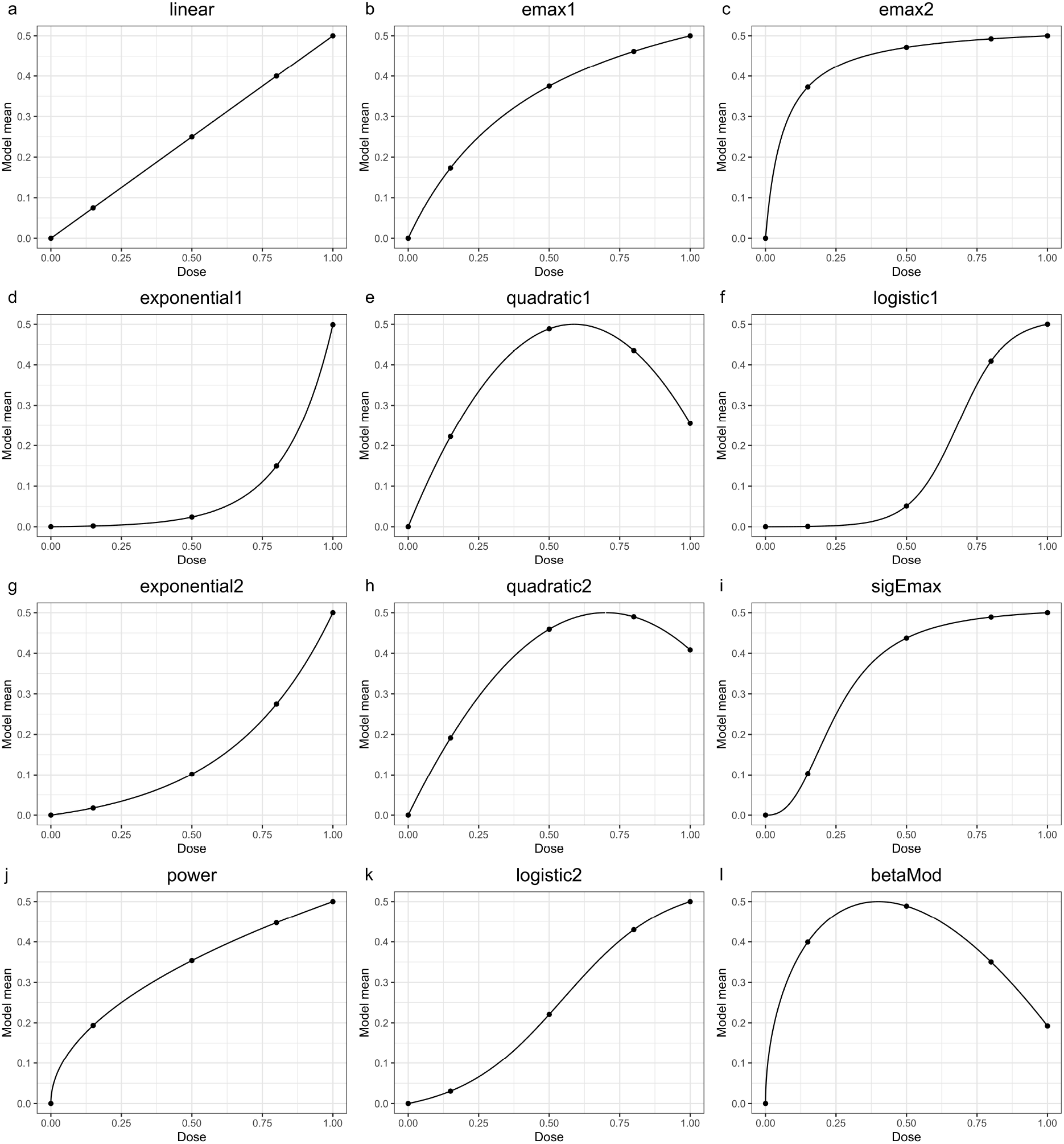
Dose-response shapes selected for the true dose-response model.

For each simulated trial, we run LiMAP-curvature to establish PoC and find the target doses of interest with a standard deviation of *τ* ∈ {1, 3, 5} for the hyperprior *HN*(*τ* ^2^). To benchmark LiMAP-curvature against smoothing spline and MCP-Mod, we run smoothing spline through the **stats** package (version 3.6.2) in R and employ the generalised cross-validation to choose the smoothing parameter for smoothing spline (Green & Silverman, 1993). We also run MCP-Mod through the **DoseFinding** package (version 1.0-4) in R, where we specify a fixed set of candidate models made up of linear, emax1, emax2, exponential1, quadratic and logistic1 (top six figures of Figure 1).

### 3.2. Simulation results

To evaluate the performance of LiMAP-curvature, smoothing spline and MCP-Mod, we plot receiver operating characteristic (ROC) curves for all true dose-response models for sample size 40 in Figure 2. The ROC curve is produced by plotting the true positive rate against the false positive rate across a range of critical values. The closer the ROC curve approaches the top left corner, the better the method performs overall. See Supplemental Material, Figures S1-S5 for ROC curves for all other sample sizes. We also summarise their powers at 5% and 10% type I error rates in Supplemental Material, Table S2 and S3, respectively.

**Figure 2:**
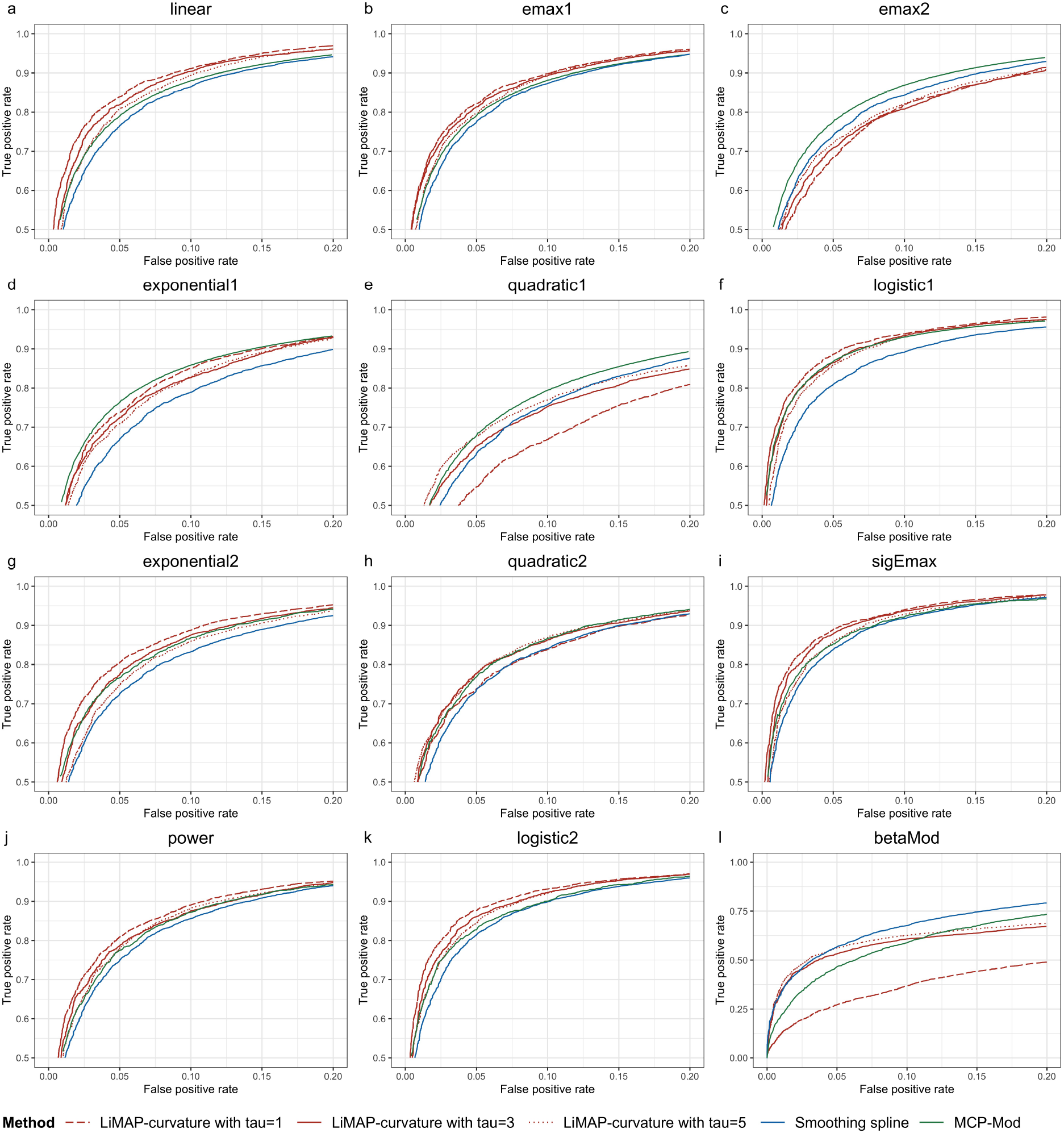
ROC curves of LiMAP-curvature, smoothing spline and MCP-Mod across different true dose-response models with the sample size of 40 patients per arm. The ROC curves of MCP-Mod in (a)-(f) are produced with the true dose-response model not included in the candidate model set, and the ROC curves of MCP-Mod in (g)-(l) are produced with the true dose-response model not included in the candidate model set.

As illustrated in Figure 2, we find that LiMAP-curvature has better performance when the true dose-response relationship is not dramatically curved, e.g. linear, emax1, logistic1, logistic2 and sigEmax, achieving over 80% power at a type I error rate of 5%. Choosing an appropriate *τ* can further improve the performance of LiMAP-curvature. More specifically, LiMAP-curvature achieves better performance with a larger *τ* for the true dose-response curve that is more curved, e.g. quadratic1 and beta models. Otherwise, e.g. in logistic1 and power models, a smaller *τ* performs better. How to select an appropriate *τ* in practice will be discussed in Section 4.

To benchmark the performance of LiMAP-curvature, smoothing spline and MCP-Mod, we consider two cases:

1. the true dose-response curve is one of the candidate models in MCP-Mod,
2. the true dose-response curve is not one of the candidate models in MCP-Mod.

Figures 2(a-f) compare the performance of LiMAP-curvature, smoothing spline and MCP-Mod for the first case, which is a fairly rare situation in practice. The resulting ROC curves are in favour of MCP-Mod as expected due to its utilisation of exact population parameter estimates in its candidate model set, but LiMAP-curvature with *τ* = 3 achieves comparable performance. We note that for the true dose-response curve that is not dramatically curved, e.g. linear, emax1 and logistic1, *τ* = 1 yields better performance than when *τ* = 3, resulting in a power gain of around 2-9% compared to MCP-Mod and around 8-11% compared to smoothing spline, all at 5% type I error rate.

Figures 2(g-l) compare the performance of LiMAP-curvature, smoothing spline and MCP-Mod for the second case. This is the situation we expect to encounter in practice. We see that with *τ* = 3, LiMAP-curvature uniformly outperforms MCP-Mod, especially when the true dose-response model drastically deviates from the candidate model set in MCP-Mod such as sigEmax and beta models. In the latter case, LiMAP-curvature has a power gain of approximately 5-13% compared to MCP-Mod at a type I error rate of 5%. Some true dose-response models such as exponential2, quadratic2 and power models, are well captured by the candidate model set of MCP-Mod so that we see the similar performance of LiMAP-curvature and MCP-Mod. Even with an inappropriate choice of *τ* , LiMAP-curvature is still able to achieve significantly better performance, e.g. a power gained by approximately 2-3% over MCP-Mod at a 5% type I error rate. Moreover, compared to the smoothing spline, LiMAP-curvature consistently outperforms it, except for the beta model. LiMAP-curvature demonstrates a power gain of about 3-9% over smoothing spline at a 5% type I error rate.

We also evaluate and compare the performance in estimating the dose-response curve and target doses of interest, known as the minimum effective dose (MED). See Supplemental Material, Figures S6-S11 for the dose-response curves estimated using LiMAP-curvature, smoothing spline and MCP-Mod for various true dose-response models and sample sizes. The corresponding results for MED estimation are summarised in Supplemental Material, Table S4.

## 4. Discussion

In this work, we have introduced LiMAP-curvature, a novel Bayesian approach for establishing PoC and estimating the dose-response curve alongside target doses of interest in Phase II trials. LiMAP-curvature is “model-free”, in the sense that it does not require pre-specification of candidate dose-response models, which can influence the performance of MCP-Mod. It is built on a Bayesian hierarchical framework incorporating prior information on the *L*^2^-total curvature of the dose-response curve.

We have shown through simulations that LiMAP-curvature has performance comparable to that of MCP-Mod in establishing PoC and estimating MED when the true dose-response model is included in the candidate model set of MCP-Mod, which is fairly rare in practice. When the true dose-response model deviates from the candidate model set of MCP-Mod, LiMAP-curvature has been demonstrated to outperform MCP-Mod. Furthermore, we note that LiMAP-curvature also outperforms smoothing spline in terms of power to detect dose-response signals in most cases. This indicates the advantages of LiMAP-curvature over the widely used non-parametric method of smoothing spline. The ability of LiMAP-curvature to consistently outperform both MCP-Mod and smoothing spline across various scenarios highlights its effectiveness and potential as a valuable tool in Phase II trial analysis.

To obtain optimal performance, MCP-Mod requires specialised expertise to pre-specify plausible candidate models and model parameters, but the knowledge of the agent/compounds being studied is usually limited. Compared to MCP-Mod, the only requirement for pre-specification in LiMAP-curvature is the standard deviation *τ* for the hyperprior *γ* ∼ *HN*(*τ* ^2^), which encodes prior knowledge of how far the dose-response curve is from a straight line. With additional simulations with varying values of the standard deviation *τ* (see Supplemental Material, Figure S12 and Table S5), we recommend choosing

1. *τ* ∈ [2, 4] if our prior knowledge is poor,
2. *τ* < 2 if we are confident that the curvature of the dose-response curve is weak,
3. *τ* > 4 if we are confident that the curvature of the dose-response curve is strong.

A number of relevant issues for LiMAP-curvature deserve further research. One such extension involves integrating the sigmoid Emax model as the default response function within the LiMAP-curvature framework. The sigmoid Emax model is noted for its ability to approximate most common monotonic dose-response relationships, providing a robust framework for modelling complex dose-response relationships. However, the challenge lies in selecting appropriate prior distributions for the additional parameters introduced by the sigmoid Emax model, which requires careful consideration to ensure accurate and reliable dose-response estimation. Another important extension involves adapting LiMAP-curvature to handle a wider variety of endpoints and trial designs. The current version of LiMAP-curvature is limited to the analysis Phase II dose-finding trials with continuous endpoints, i.e. a single normally distributed homoscedastic response measured at the end of the trial. To expand its applicability, extensions to other common types of endpoints (e.g. binary, counts and survival endpoints) and trials (e.g. longitudinal dose-finding trials) require further investigation. Other directions of future research include the investigation of trial designs tailored to LiMAP-curvature and the development of statistical software implementing LiMAP-curvature.

## Data Availability

All data produced in the present work are simulated.

## Disclaimer

The opinions expressed in this paper are solely those of the authors and not those of their affiliations. The authors’ affiliations do not guarantee the accuracy or reliability of the information provided herein.

