## Supplemental_Material for "LiMAP-curvature: a simple model-free approach for analysing dose-finding studies"

| Model | Functional form $f(d, \theta)$ | Model specification |
|---|---|---|
| linear | $E_0 + \delta d$ | $E_0 = 0,\ \delta = 0.5$ |
| emax1 | $E_0 + E_{\max} d/(ED_{50} + d)$ | $E_0 = 0,\ E_{\max} = 0.75,\ ED_{50} = 0.5$ |
| emax2 | | $E_0 = 0,\ E_{\max} = 0.5321,\ ED_{50} = 0.0642$ |
| exponential1 | $E_0 + E_1(\exp(d/\delta) - 1)$ | $E_0 = 0,\ E_1 = 0.00125,\ \delta = 0.1669$ |
| exponential2 | | $E_0 = 0,\ E_1 = 0.3492,\ \delta = 0.3664$ |
| quadratic1 | $E_0 + \beta_1 d + \beta_2 d^2$ | $E_0 = 0,\ \beta_1 = 1.70,\ \beta_2 = -1.445$ |
| quadratic2 | | $E_0 = 0,\ \beta_1 = 1.4286,\ \beta_2 = -1.0204$ |
| logistic1 | $E_0 + E_{\max}/(1 + \exp((ED_{50} - d)/\delta))$ | $E_0 = -0.0001,\ E_{\max} = 0.5116,\ ED_{50} = 0.6839,\ \delta = 0.0837$ |
| logistic2 | | $E_0 = -0.0264,\ E_{\max} = 0.5701,\ ED_{50} = 0.5485,\ \delta = 0.1813$ |
| power | $E_0 + E_{\max} d^h$ | $E_0 = 0,\ E_{\max} = 0.5,\ h = 0.5$ |
| sigmoid Emax(sigEmax) | $E_0 + E_{\max} d^h/(ED_{50}^h + d^h)$ | $E_0 = 0,\ E_{\max} = 0.5146,\ ED_{50} = 0.2561,\ h = 2.5922$ |
| beta model (betaMod) | $E_0 + E_{\max} B(\delta_1, \delta_2)(d/scal)^{\delta_1}(1 - d/scal)^{\delta_2}$ | $E_0 = 0,\ E_{\max} = 0.5,\ \delta_1 = 0.3518,\ \delta_2 = 1.0554,\ scal = 1.2$ |

Table S1: Model specifications for the dose-response shapes selected for the true underlying dose-response model. For the beta model, $B(\delta_1, \delta_2) = (\delta_1 + \delta_2)^{\delta_1 + \delta_2}/(\delta_1^{\delta_1} \delta_2^{\delta_2})$.

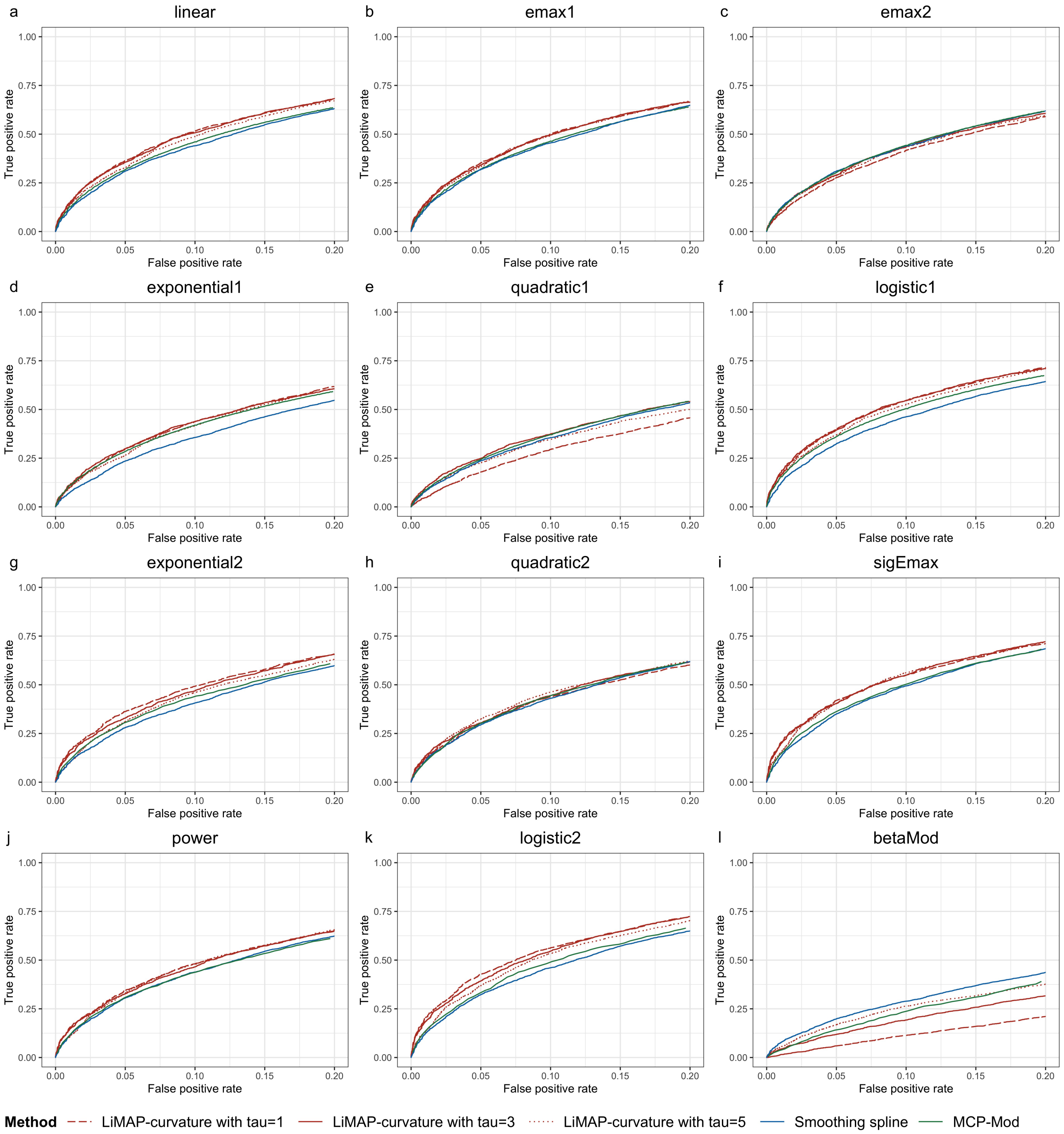


Figure S1: ROC curves of LiMAP-curvature, smoothing spline and MCP-Mod across different true underlying dose-response models with the sample size of 10 patients per arm. The ROC curves of MCP-Mod in (a)-(f) are produced with the true underlying dose-response model not included in the candidate model set, and the ROC curves of MCP-Mod in (g)-(l) are produced with the true underlying dose-response model not included in the candidate model set.

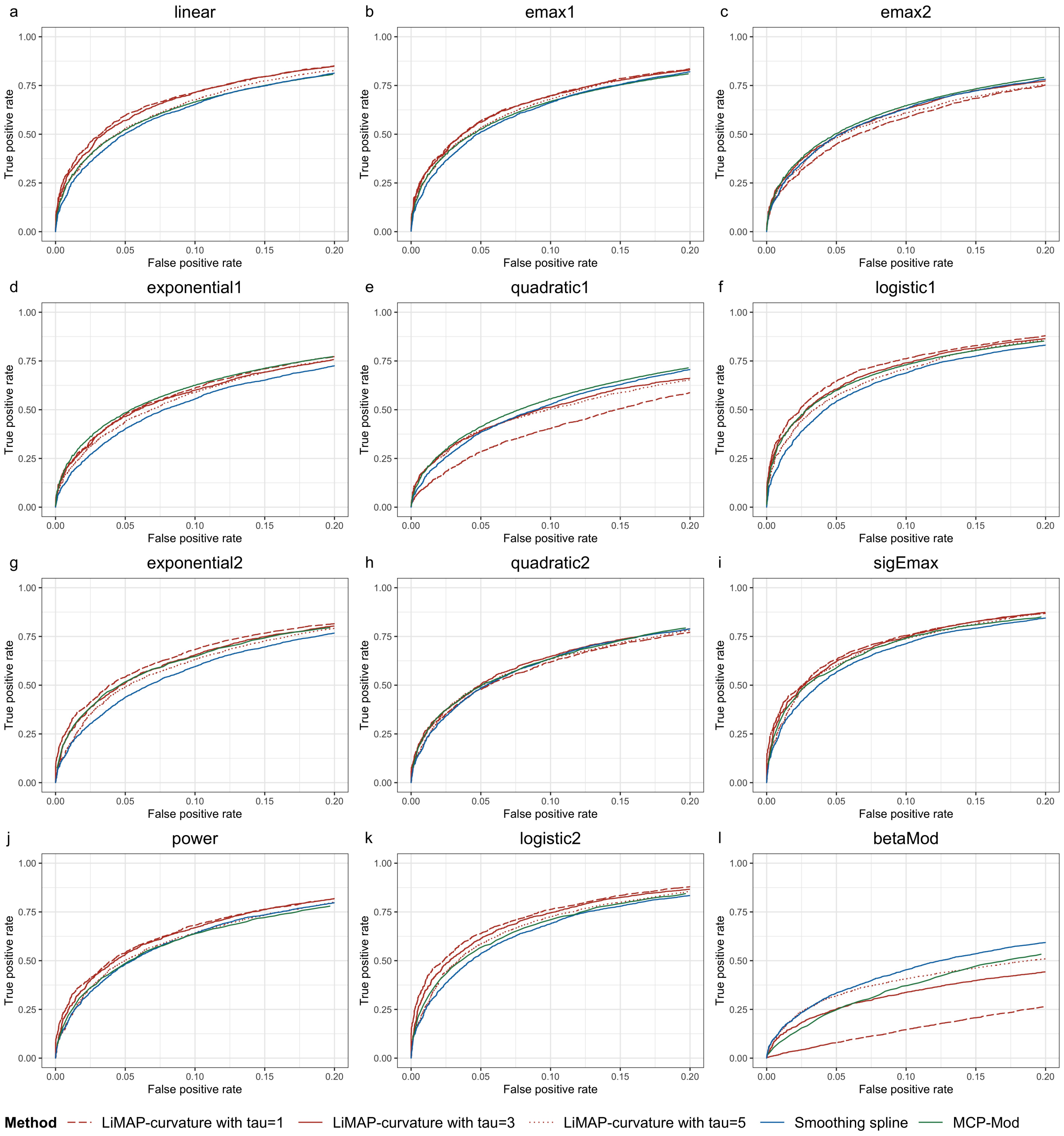


Figure S2: ROC curves of LiMAP-curvature, smoothing spline and MCP-Mod across different true underlying dose-response models with the sample size of 20 patients per arm. The ROC curves of MCP-Mod in (a)-(f) are produced with the true underlying dose-response model not included in the candidate model set, and the ROC curves of MCP-Mod in (g)-(l) are produced with the true underlying dose-response model not included in the candidate model set.

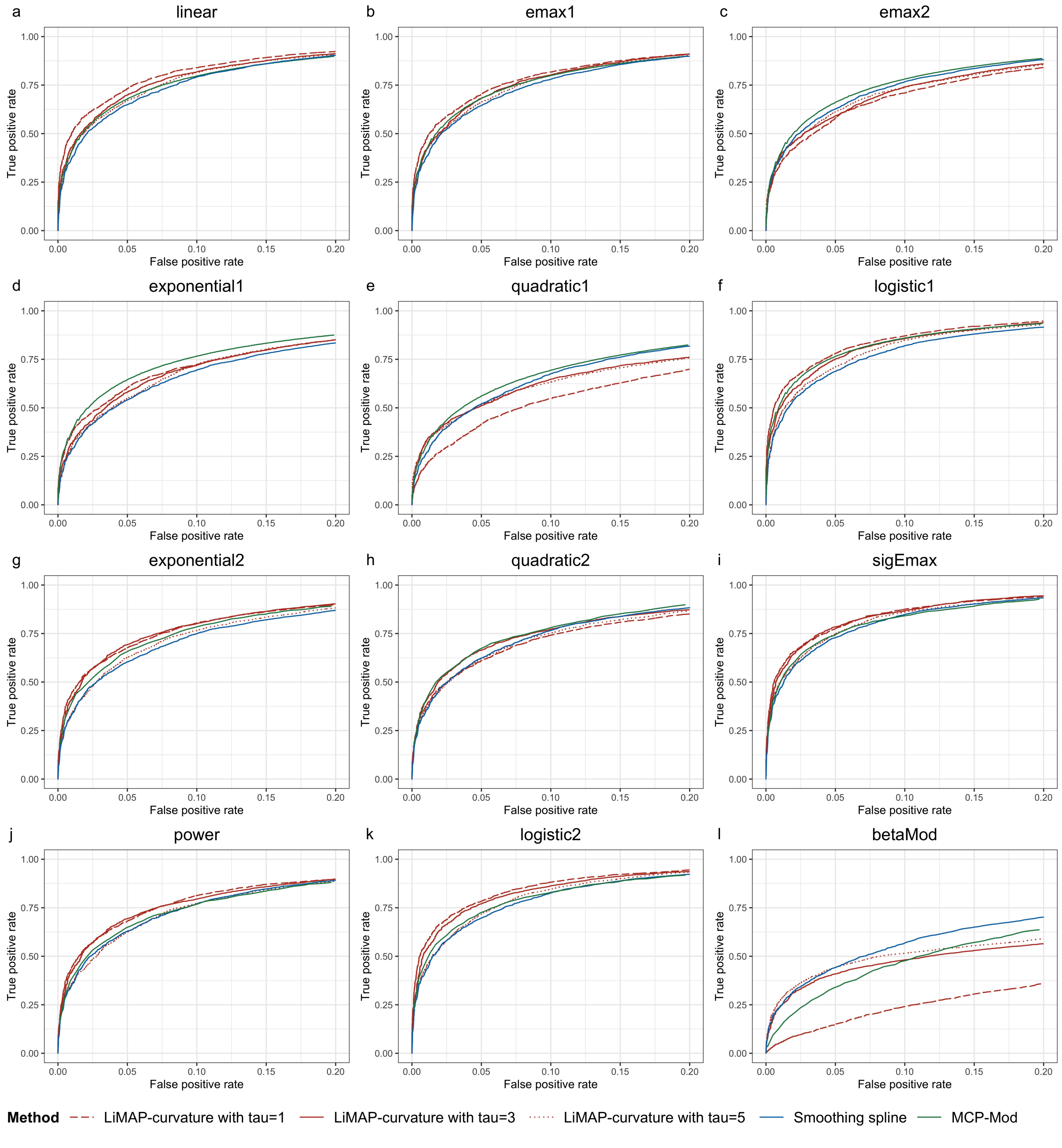


Figure S3: ROC curves of LiMAP-curvature, smoothing spline and MCP-Mod across different true underlying dose-response models with the sample size of 30 patients per arm. The ROC curves of MCP-Mod in (a)-(f) are produced with the true underlying dose-response model not included in the candidate model set, and the ROC curves of MCP-Mod in (g)-(l) are produced with the true underlying dose-response model not included in the candidate model set.

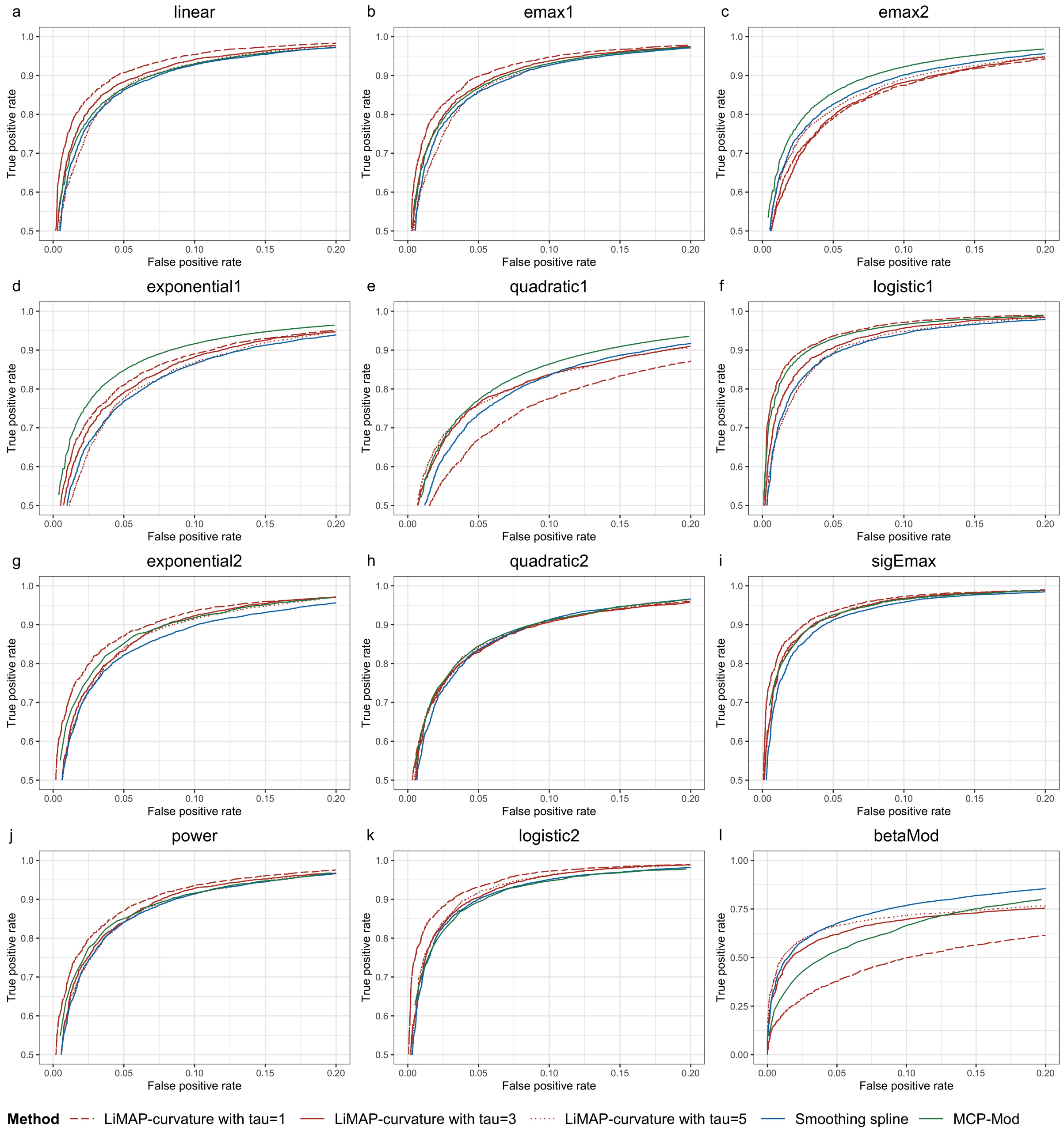


Figure S4: ROC curves of LiMAP-curvature, smoothing spline and MCP-Mod across different true underlying dose-response models with the sample size of 50 patients per arm. The ROC curves of MCP-Mod in (a)-(f) are produced with the true underlying dose-response model not included in the candidate model set, and the ROC curves of MCP-Mod in (g)-(l) are produced with the true underlying dose-response model not included in the candidate model set.

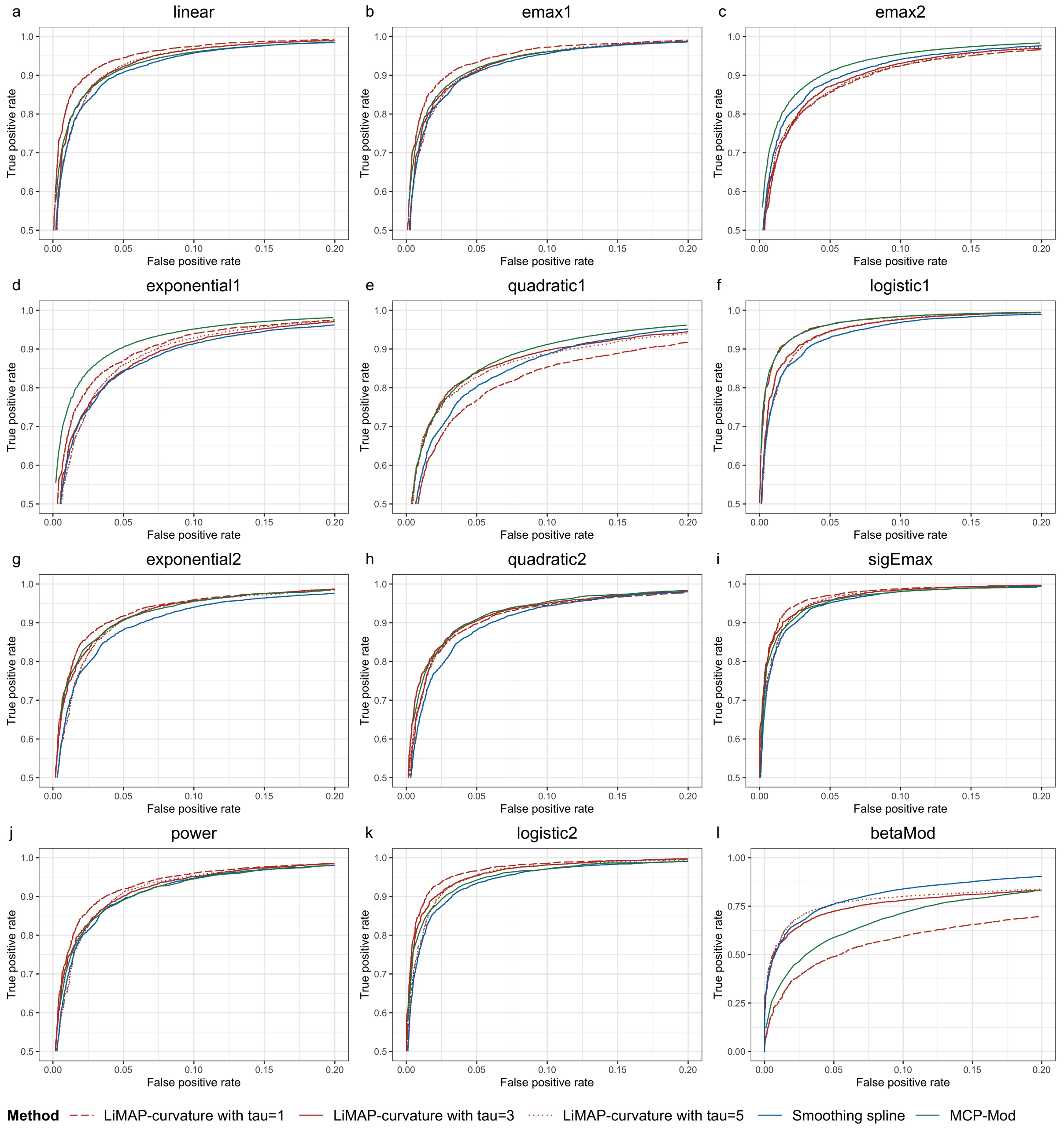


Figure S5: ROC curves of LiMAP-curvature, smoothing spline and MCP-Mod across different true underlying dose-response models with the sample size of 60 patients per arm. The ROC curves of MCP-Mod in (a)-(f) are produced with the true underlying dose-response model not included in the candidate model set, and the ROC curves of MCP-Mod in (g)-(l) are produced with the true underlying dose-response model not included in the candidate model set.

| Sample size | Model | LiMAP-curvature | | | Smoothing spline | MCP-Mod |
|---|---|---|---|---|---|---|
| | | $\tau = 1$ | $\tau = 3$ | $\tau = 5$ | | |
| $N = 10$ | linear | 0.376 | 0.372 | 0.338 | 0.310 | 0.317 |
| | emax1 | 0.360 | 0.370 | 0.327 | 0.317 | 0.319 |
| | emax2 | 0.280 | 0.302 | 0.290 | 0.309 | 0.302 |
| | exponential1 | 0.296 | 0.304 | 0.266 | 0.234 | 0.284 |
| | quadratic1 | 0.182 | 0.233 | 0.244 | 0.236 | 0.243 |
| | logistic1 | 0.418 | 0.411 | 0.357 | 0.322 | 0.359 |
| | exponential2 | 0.361 | 0.336 | 0.300 | 0.279 | 0.299 |
| | quadratic2 | 0.306 | 0.324 | 0.318 | 0.294 | 0.282 |
| | sigEmax | 0.411 | 0.414 | 0.388 | 0.349 | 0.347 |
| | power | 0.355 | 0.331 | 0.313 | 0.197 | 0.307 |
| | logistic2 | 0.429 | 0.396 | 0.374 | 0.322 | 0.342 |
| | betaMod | 0.066 | 0.121 | 0.166 | 0.197 | 0.146 |
| $N = 20$ | linear | 0.611 | 0.586 | 0.544 | 0.503 | 0.523 |
| | emax1 | 0.582 | 0.553 | 0.548 | 0.511 | 0.525 |
| | emax2 | 0.464 | 0.473 | 0.491 | 0.492 | 0.503 |
| | exponential1 | 0.497 | 0.465 | 0.443 | 0.401 | 0.484 |
| | quadratic1 | 0.446 | 0.519 | 0.524 | 0.384 | 0.413 |
| | logistic1 | 0.656 | 0.629 | 0.563 | 0.542 | 0.598 |
| | exponential2 | 0.546 | 0.541 | 0.494 | 0.438 | 0.488 |
| | quadratic2 | 0.498 | 0.531 | 0.500 | 0.485 | 0.521 |
| | sigEmax | 0.628 | 0.634 | 0.624 | 0.566 | 0.575 |
| | power | 0.558 | 0.549 | 0.498 | 0.481 | 0.501 |
| | logistic2 | 0.658 | 0.624 | 0.585 | 0.536 | 0.560 |
| | betaMod | 0.082 | 0.252 | 0.306 | 0.334 | 0.250 |
| $N = 30$ | linear | 0.742 | 0.728 | 0.659 | 0.649 | 0.681 |
| | emax1 | 0.719 | 0.699 | 0.656 | 0.645 | 0.683 |
| | emax2 | 0.589 | 0.591 | 0.634 | 0.626 | 0.662 |
| | exponential1 | 0.622 | 0.604 | 0.585 | 0.540 | 0.646 |

**Table S2 continued from previous page**

| | | | | | | |
|---|---|---|---|---|---|---|
| | quadratic1 | 0.537 | 0.651 | 0.663 | 0.520 | 0.562 |
| | logistic1 | 0.781 | 0.774 | 0.740 | 0.688 | 0.766 |
| | exponential2 | 0.697 | 0.664 | 0.622 | 0.603 | 0.663 |
| | quadratic2 | 0.621 | 0.650 | 0.638 | 0.623 | 0.660 |
| | sigEmax | 0.779 | 0.766 | 0.748 | 0.728 | 0.731 |
| | power | 0.685 | 0.661 | 0.640 | 0.629 | 0.657 |
| | logistic2 | 0.780 | 0.754 | 0.717 | 0.696 | 0.711 |
| | betaMod | 0.156 | 0.404 | 0.439 | 0.442 | 0.332 |
| $N = 40$ | linear | 0.839 | 0.819 | 0.786 | 0.755 | 0.788 |
| | emax1 | 0.884 | 0.799 | 0.777 | 0.768 | 0.791 |
| | emax2 | 0.685 | 0.710 | 0.722 | 0.738 | 0.774 |
| | exponential1 | 0.724 | 0.709 | 0.698 | 0.655 | 0.763 |
| | quadratic1 | 0.547 | 0.652 | 0.676 | 0.622 | 0.678 |
| | logistic1 | 0.886 | 0.851 | 0.827 | 0.795 | 0.867 |
| | exponential2 | 0.804 | 0.774 | 0.747 | 0.711 | 0.777 |
| | quadratic2 | 0.738 | 0.778 | 0.761 | 0.727 | 0.752 |
| | sigEmax | 0.889 | 0.878 | 0.858 | 0.832 | 0.830 |
| | power | 0.809 | 0.786 | 0.760 | 0.740 | 0.779 |
| | logistic2 | 0.891 | 0.863 | 0.843 | 0.812 | 0.826 |
| | betaMod | 0.271 | 0.532 | 0.562 | 0.555 | 0.424 |
| $N = 50$ | linear | 0.913 | 0.868 | 0.864 | 0.862 | 0.867 |
| | emax1 | 0.888 | 0.867 | 0.863 | 0.857 | 0.868 |
| | emax2 | 0.773 | 0.790 | 0.797 | 0.825 | 0.856 |
| | exponential1 | 0.803 | 0.786 | 0.773 | 0.767 | 0.848 |
| | quadratic1 | 0.651 | 0.757 | 0.763 | 0.734 | 0.773 |
| | logistic1 | 0.931 | 0.911 | 0.898 | 0.890 | 0.929 |
| | exponential2 | 0.873 | 0.832 | 0.823 | 0.821 | 0.870 |
| | quadratic2 | 0.842 | 0.830 | 0.836 | 0.836 | 0.853 |
| | sigEmax | 0.942 | 0.920 | 0.916 | 0.912 | 0.911 |

**Table S2 continued from previous page**

| | | | | | | |
|---|---|---|---|---|---|---|
| | power | 0.880 | 0.834 | 0.830 | 0.841 | 0.838 |
| | logistic2 | 0.939 | 0.906 | 0.902 | 0.900 | 0.900 |
| | betaMod | 0.384 | 0.626 | 0.665 | 0.674 | 0.525 |
| $N = 60$ | linear | 0.938 | 0.936 | 0.913 | 0.906 | 0.916 |
| | emax1 | 0.933 | 0.923 | 0.916 | 0.902 | 0.917 |
| | emax2 | 0.923 | 0.937 | 0.927 | 0.890 | 0.909 |
| | exponential1 | 0.873 | 0.857 | 0.855 | 0.830 | 0.904 |
| | quadratic1 | 0.759 | 0.839 | 0.841 | 0.798 | 0.841 |
| | logistic1 | 0.966 | 0.948 | 0.946 | 0.930 | 0.963 |
| | exponential2 | 0.917 | 0.891 | 0.888 | 0.872 | 0.903 |
| | quadratic2 | 0.900 | 0.902 | 0.885 | 0.877 | 0.903 |
| | sigEmax | 0.966 | 0.958 | 0.950 | 0.951 | 0.946 |
| | power | 0.919 | 0.896 | 0.887 | 0.740 | 0.898 |
| | logistic2 | 0.960 | 0.947 | 0.948 | 0.932 | 0.943 |
| | betaMod | 0.498 | 0.721 | 0.755 | 0.740 | 0.586 |

Table S2: Power values of LiMAP-curvature, smoothing spline and MCP-Mod to test the PoC across different true underlying dose-response models and sample sizes by controlling the type I error rate at 5%.

| Sample size | Model | LiMAP-curvature | | | Smoothing spline | MCP-Mod |
|---|---|---|---|---|---|---|
| | | $\tau = 1$ | $\tau = 3$ | $\tau = 5$ | | |
| $N = 10$ | linear | 0.516 | 0.501 | 0.497 | 0.440 | 0.460 |
| | emax1 | 0.505 | 0.490 | 0.494 | 0.455 | 0.463 |
| | emax2 | 0.415 | 0.411 | 0.442 | 0.439 | 0.442 |
| | exponential1 | 0.438 | 0.407 | 0.415 | 0.356 | 0.419 |
| | quadratic1 | 0.295 | 0.332 | 0.379 | 0.355 | 0.370 |
| | logistic1 | 0.558 | 0.532 | 0.525 | 0.462 | 0.504 |
| | exponential2 | 0.493 | 0.453 | 0.463 | 0.407 | 0.435 |
| | quadratic2 | 0.438 | 0.437 | 0.451 | 0.430 | 0.432 |
| | sigEmax | 0.543 | 0.534 | 0.550 | 0.494 | 0.507 |
| | power | 0.474 | 0.469 | 0.475 | 0.438 | 0.428 |
| | logistic2 | 0.544 | 0.520 | 0.529 | 0.460 | 0.489 |
| | betaMod | 0.113 | 0.191 | 0.252 | 0.289 | 0.257 |
| $N = 20$ | linear | 0.775 | 0.706 | 0.699 | 0.652 | 0.663 |
| | emax1 | 0.718 | 0.695 | 0.693 | 0.661 | 0.667 |
| | emax2 | 0.605 | 0.619 | 0.623 | 0.630 | 0.645 |
| | exponential1 | 0.638 | 0.611 | 0.588 | 0.554 | 0.624 |
| | quadratic1 | 0.446 | 0.519 | 0.524 | 0.526 | 0.554 |
| | logistic1 | 0.778 | 0.755 | 0.712 | 0.687 | 0.729 |
| | exponential2 | 0.680 | 0.675 | 0.653 | 0.595 | 0.638 |
| | quadratic2 | 0.628 | 0.649 | 0.640 | 0.635 | 0.657 |
| | sigEmax | 0.753 | 0.756 | 0.759 | 0.712 | 0.715 |
| | power | 0.685 | 0.679 | 0.655 | 0.640 | 0.646 |
| | logistic2 | 0.775 | 0.747 | 0.727 | 0.688 | 0.717 |
| | betaMod | 0.158 | 0.331 | 0.404 | 0.453 | 0.359 |
| $N = 30$ | linear | 0.845 | 0.822 | 0.806 | 0.792 | 0.797 |
| | emax1 | 0.825 | 0.820 | 0.799 | 0.780 | 0.799 |
| | emax2 | 0.716 | 0.738 | 0.734 | 0.767 | 0.781 |
| | exponential1 | 0.732 | 0.729 | 0.713 | 0.694 | 0.766 |

**Table S3 continued from previous page**

| | | | | | | |
|---|---|---|---|---|---|---|
| | quadratic1 | 0.550 | 0.632 | 0.652 | 0.676 | 0.693 |
| | logistic1 | 0.884 | 0.861 | 0.842 | 0.820 | 0.859 |
| | exponential2 | 0.809 | 0.781 | 0.770 | 0.749 | 0.779 |
| | quadratic2 | 0.758 | 0.759 | 0.768 | 0.765 | 0.784 |
| | sigEmax | 0.873 | 0.858 | 0.866 | 0.848 | 0.839 |
| | power | 0.780 | 0.778 | 0.786 | 0.770 | 0.776 |
| | logistic2 | 0.875 | 0.860 | 0.855 | 0.826 | 0.827 |
| | betaMod | 0.233 | 0.461 | 0.520 | 0.568 | 0.471 |
| $N = 40$ | linear | 0.911 | 0.904 | 0.885 | 0.866 | 0.880 |
| | emax1 | 0.892 | 0.890 | 0.880 | 0.875 | 0.881 |
| | emax2 | 0.819 | 0.821 | 0.808 | 0.846 | 0.868 |
| | exponential1 | 0.826 | 0.829 | 0.823 | 0.789 | 0.858 |
| | quadratic1 | 0.668 | 0.769 | 0.752 | 0.753 | 0.793 |
| | logistic1 | 0.938 | 0.933 | 0.912 | 0.890 | 0.929 |
| | exponential2 | 0.888 | 0.875 | 0.859 | 0.890 | 0.869 |
| | quadratic2 | 0.839 | 0.863 | 0.853 | 0.845 | 0.856 |
| | sigEmax | 0.940 | 0.937 | 0.928 | 0.916 | 0.920 |
| | power | 0.891 | 0.876 | 0.869 | 0.898 | 0.851 |
| | logistic2 | 0.938 | 0.933 | 0.916 | 0.853 | 0.908 |
| | betaMod | 0.367 | 0.608 | 0.626 | 0.679 | 0.560 |
| $N = 50$ | linear | 0.957 | 0.939 | 0.932 | 0.927 | 0.929 |
| | emax1 | 0.947 | 0.934 | 0.931 | 0.926 | 0.930 |
| | emax2 | 0.872 | 0.883 | 0.879 | 0.901 | 0.921 |
| | exponential1 | 0.893 | 0.885 | 0.879 | 0.864 | 0.915 |
| | quadratic1 | 0.767 | 0.831 | 0.836 | 0.834 | 0.863 |
| | logistic1 | 0.969 | 0.960 | 0.952 | 0.942 | 0.965 |
| | exponential2 | 0.931 | 0.922 | 0.908 | 0.898 | 0.936 |
| | quadratic2 | 0.907 | 0.910 | 0.908 | 0.912 | 0.923 |
| | sigEmax | 0.974 | 0.964 | 0.961 | 0.957 | 0.959 |

**Table S3 continued from previous page**

| | | | | | | |
|---|---|---|---|---|---|---|
| | power | 0.935 | 0.917 | 0.911 | 0.913 | 0.928 |
| | logistic2 | 0.969 | 0.961 | 0.954 | 0.950 | 0.953 |
| | betaMod | 0.488 | 0.703 | 0.720 | 0.767 | 0.649 |
| $N = 60$ | linear | 0.972 | 0.968 | 0.963 | 0.954 | 0.960 |
| | emax1 | 0.966 | 0.968 | 0.962 | 0.955 | 0.960 |
| | emax2 | 0.923 | 0.937 | 0.927 | 0.943 | 0.955 |
| | exponential1 | 0.932 | 0.931 | 0.930 | 0.906 | 0.951 |
| | quadratic1 | 0.850 | 0.900 | 0.897 | 0.883 | 0.911 |
| | logistic1 | 0.987 | 0.982 | 0.975 | 0.967 | 0.984 |
| | exponential2 | 0.957 | 0.949 | 0.949 | 0.934 | 0.953 |
| | quadratic2 | 0.950 | 0.952 | 0.934 | 0.943 | 0.955 |
| | sigEmax | 0.986 | 0.984 | 0.979 | 0.980 | 0.977 |
| | power | 0.964 | 0.955 | 0.948 | 0.945 | 0.943 |
| | logistic2 | 0.984 | 0.979 | 0.980 | 0.967 | 0.973 |
| | betaMod | 0.604 | 0.781 | 0.802 | 0.825 | 0.708 |

Table S3: Power values of LiMAP-curvature, smoothing spline and MCP-Mod to test the PoC across different true underlying dose-response models and sample sizes by controlling the type I error rate at 10%.

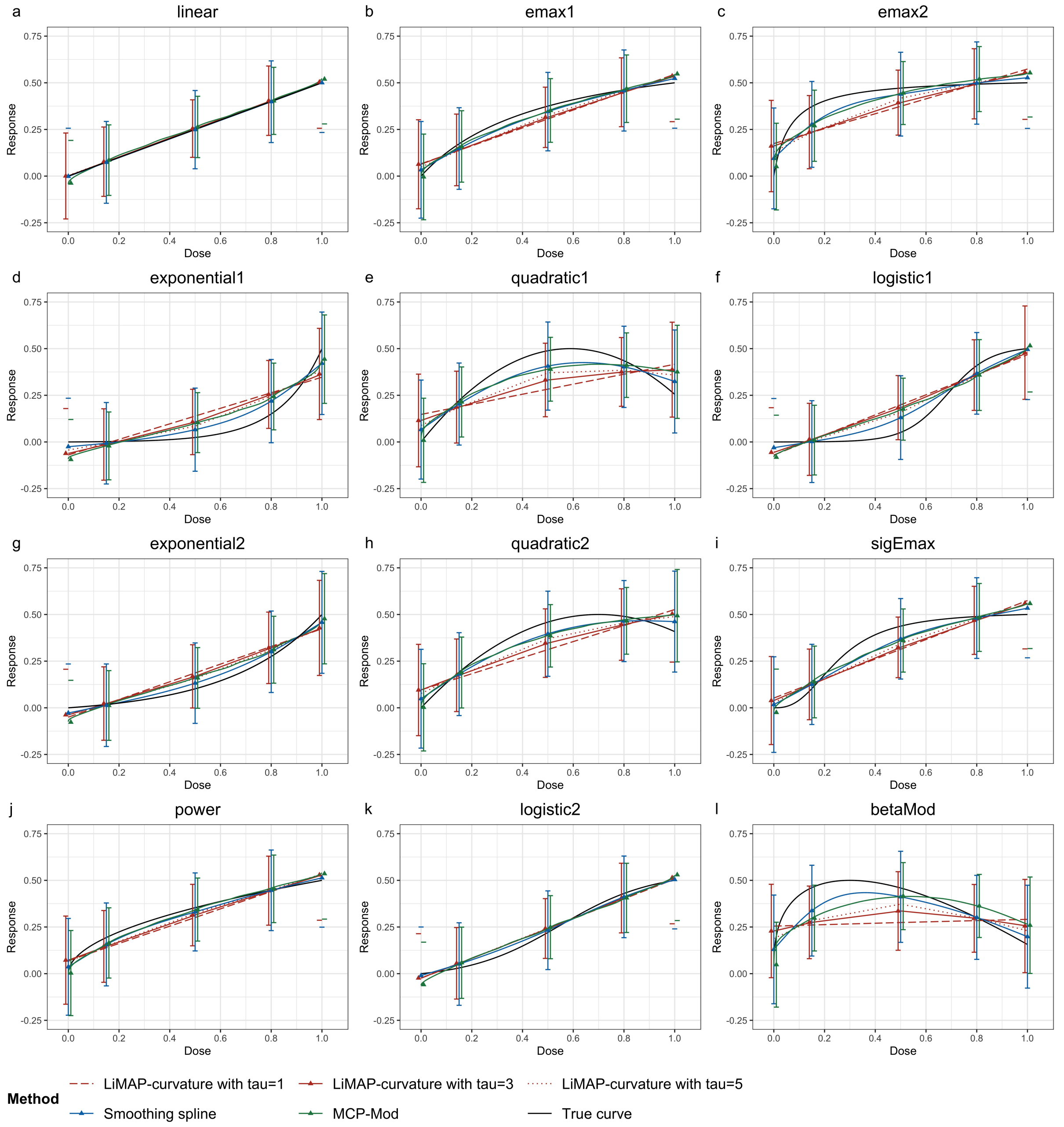


Figure S6: Dose-response curves estimated with LiMAP-curvature, smoothing spline and MCP-Mod across different true underlying dose-response models with the sample size of 10 patients per arm. The dose-response curves in (a)-(f) are produced through MCP-Mod with the true underlying dose-response model not included in the candidate model set, and the dose-response curves in (g)-(l) are produced through MCP-Mod with the true underlying dose-response model not included in the candidate model set.

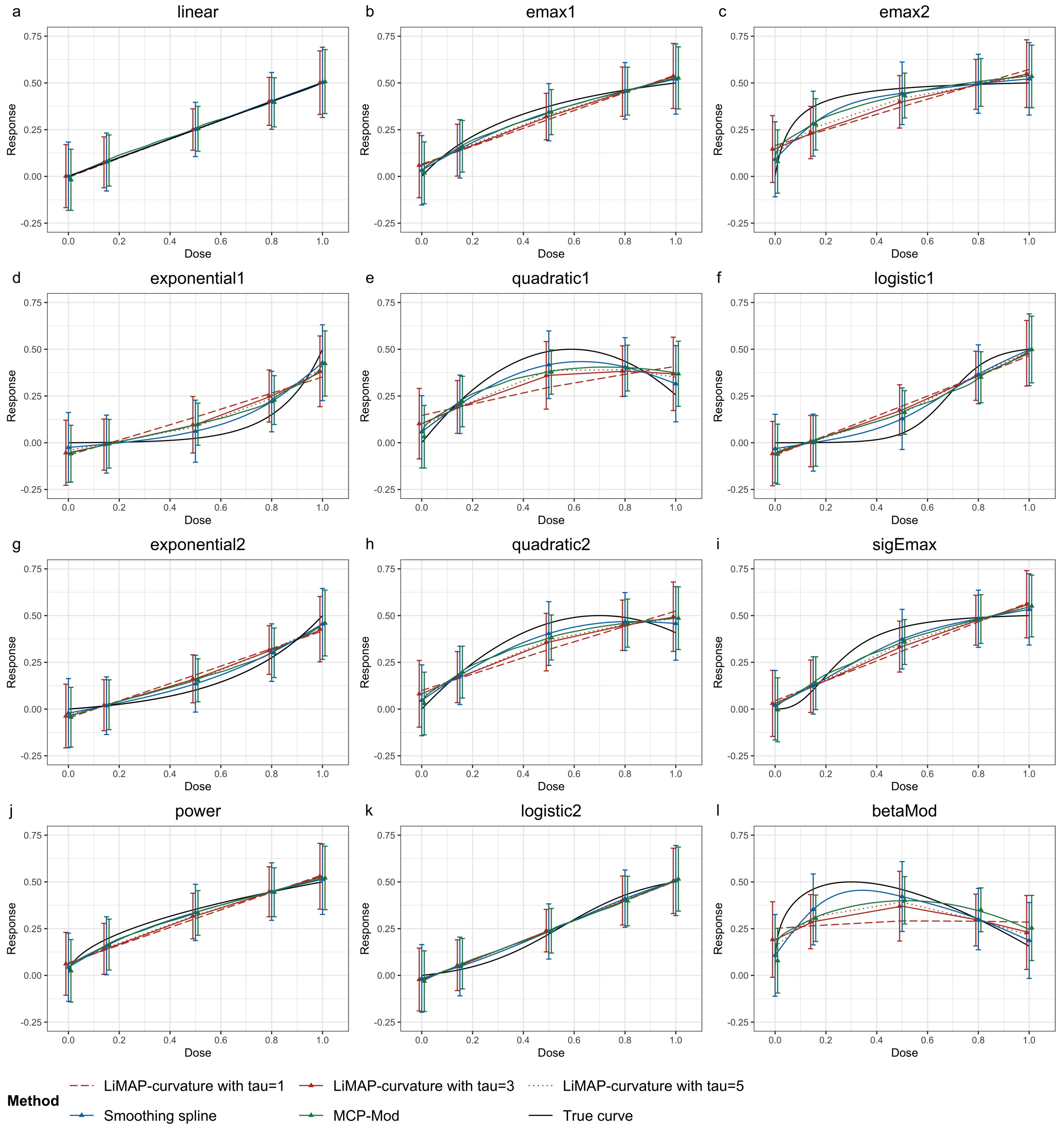


Figure S7: Dose-response curves estimated with LiMAP-curvature, smoothing spline and MCP-Mod across different true underlying dose-response models with the sample size of 20 patients per arm. The dose-response curves in (a)-(f) are produced through MCP-Mod with the true underlying dose-response model not included in the candidate model set, and the dose-response curves in (g)-(l) are produced through MCP-Mod with the true underlying dose-response model not included in the candidate model set.

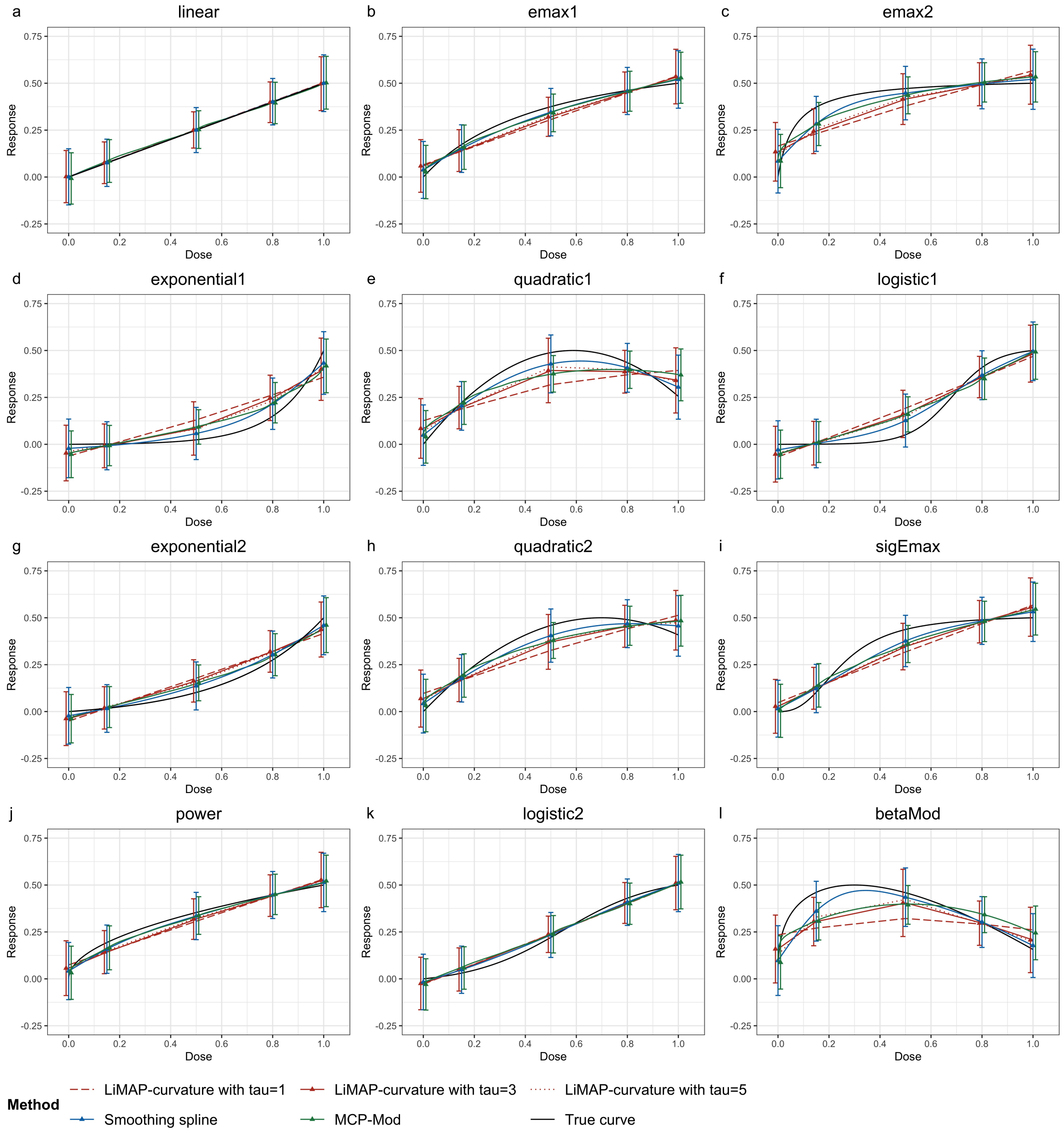


Figure S8: Dose-response curves estimated with LiMAP-curvature, smoothing spline and MCP-Mod across different true underlying dose-response models with the sample size of 30 patients per arm. The dose-response curves in (a)-(f) are produced through MCP-Mod with the true underlying dose-response model not included in the candidate model set, and the dose-response curves in (g)-(l) are produced through MCP-Mod with the true underlying dose-response model not included in the candidate model set.

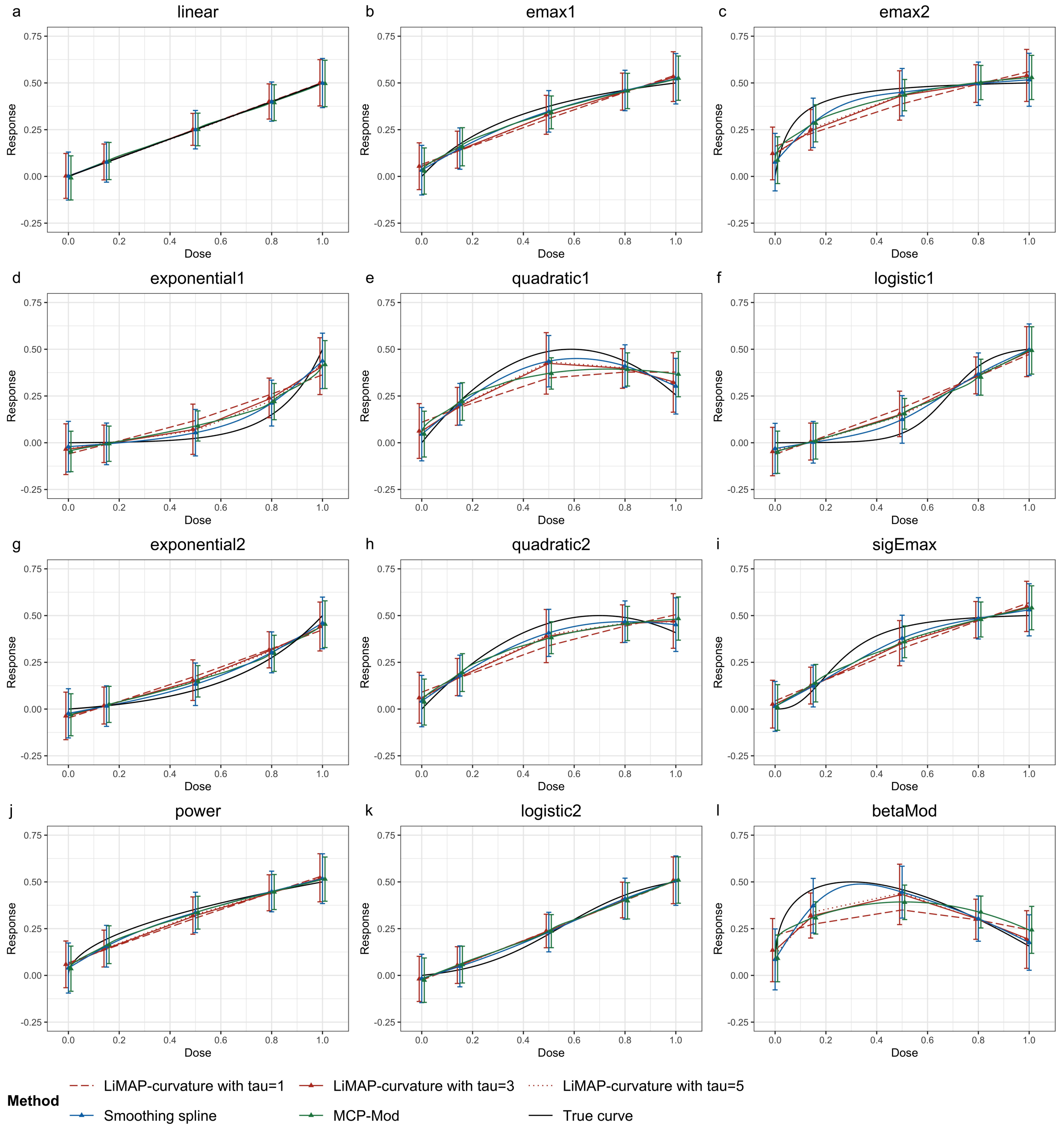


Figure S9: Dose-response curves estimated with LiMAP-curvature, smoothing spline and MCP-Mod across different true underlying dose-response models with the sample size of 40 patients per arm. The dose-response curves in (a)-(f) are produced through MCP-Mod with the true underlying dose-response model not included in the candidate model set, and the dose-response curves in (g)-(l) are produced through MCP-Mod with the true underlying dose-response model not included in the candidate model set.

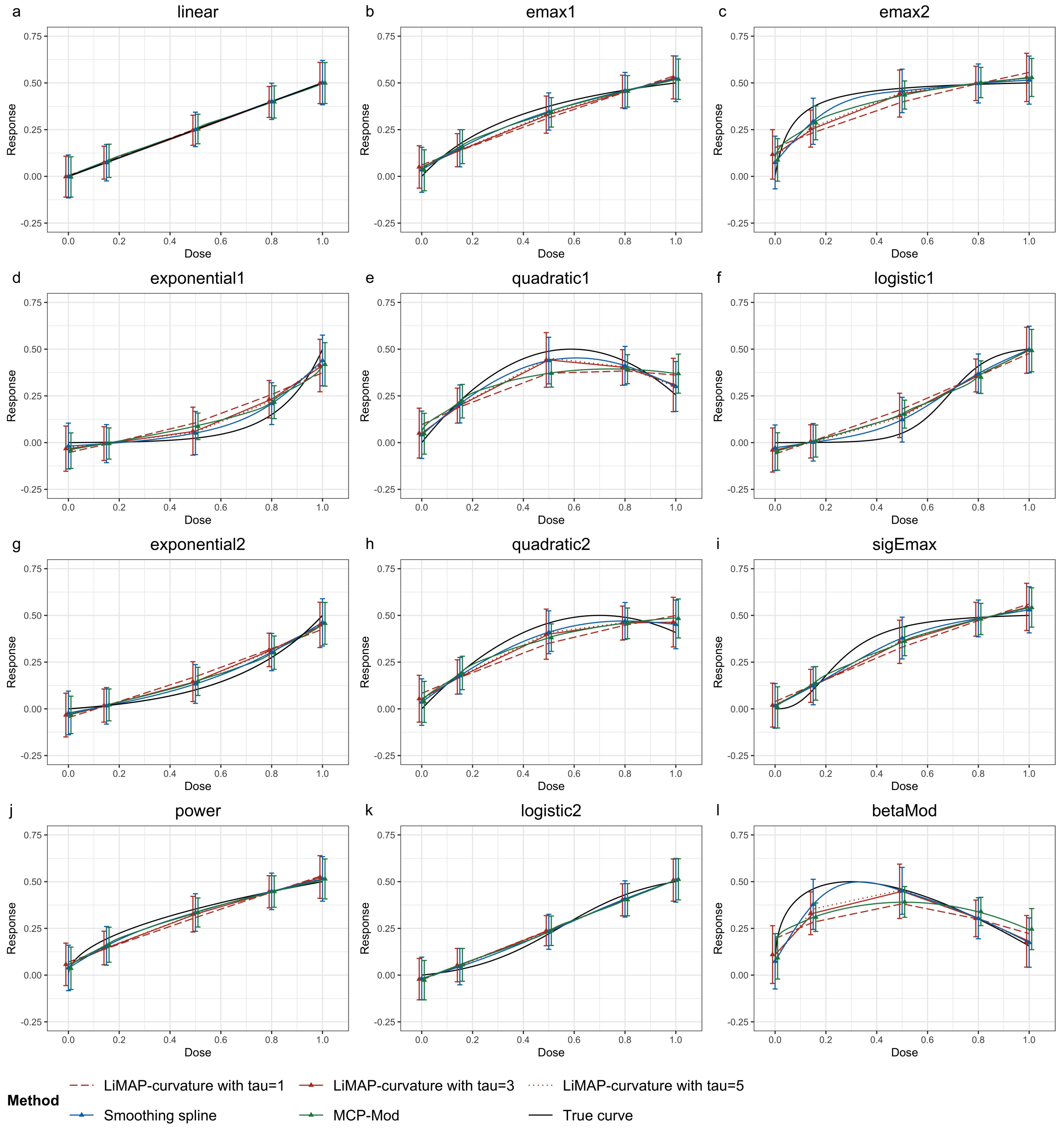


Figure S10: Dose-response curves estimated with LiMAP-curvature, smoothing spline and MCP-Mod across different true underlying dose-response models with the sample size of 50 patients per arm. The dose-response curves in (a)-(f) are produced through MCP-Mod with the true underlying dose-response model not included in the candidate model set, and the dose-response curves in (g)-(l) are produced through MCP-Mod with the true underlying dose-response model not included in the candidate model set.

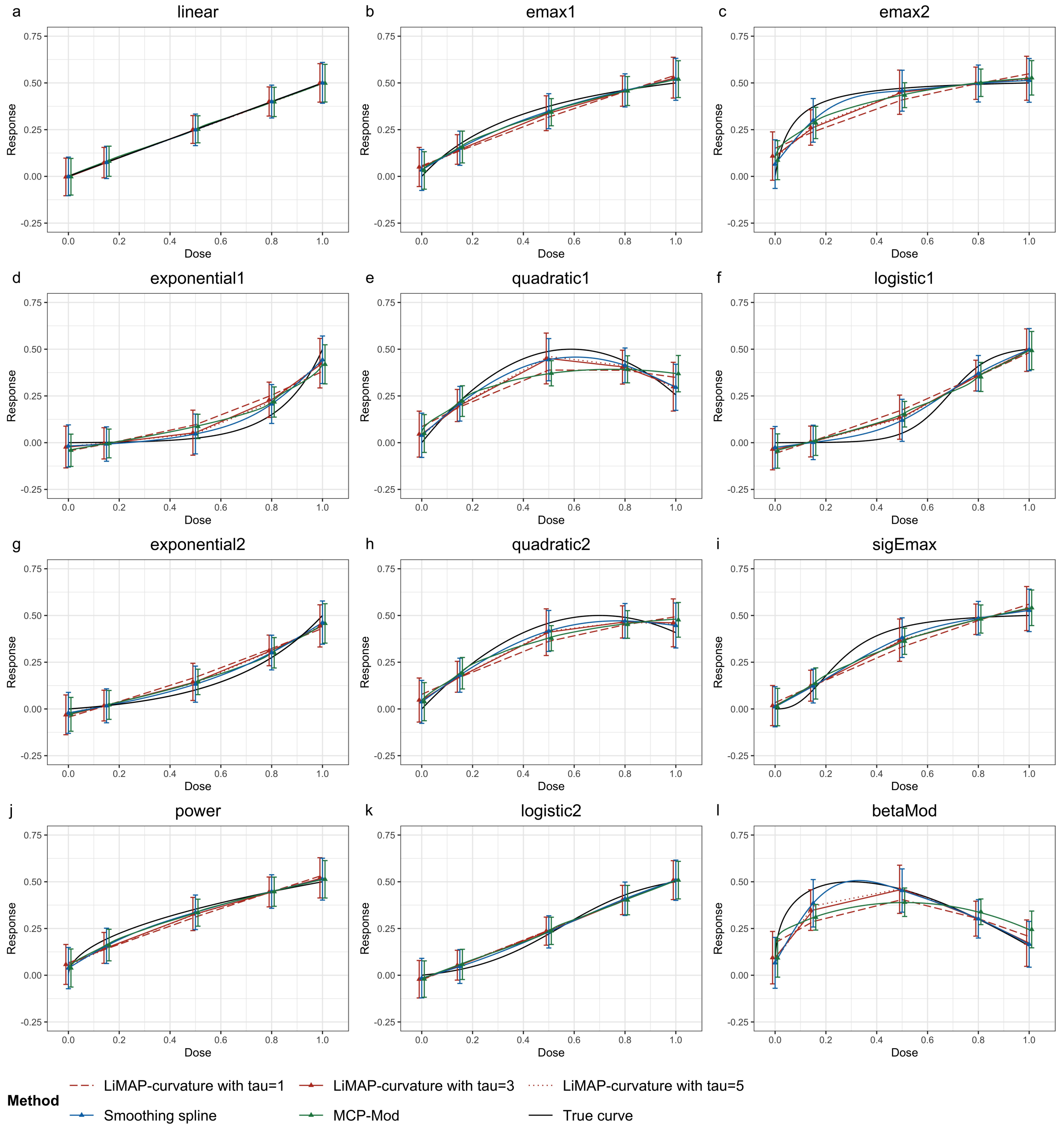


Figure S11: Dose-response curves estimated with LiMAP-curvature, smoothing spline and MCP-Mod across different true underlying dose-response models with the sample size of 60 patients per arm. The dose-response curves in (a)-(f) are produced through MCP-Mod with the true underlying dose-response model not included in the candidate model set, and the dose-response curves in (g)-(l) are produced through MCP-Mod with the true underlying dose-response model not included in the candidate model set.

| Sample size | Model | True MED | LiMAP-curvature | | | | | | Smoothing spline | | MCP-Mod | |
|---|---|---|---|---|---|---|---|---|---|---|---|---|
| | | | $\tau = 1$ | | $\tau = 3$ | | $\tau = 5$ | | | | | |
| | | | Bias | MSE | Bias | MSE | Bias | MSE | Bias | MSE | Bias | MSE |
| $N = 10$ | linear | 0.600 | -0.098 | 0.046 | -0.098 | 0.048 | -0.100 | 0.053 | -0.139 | 0.071 | -0.120 | 0.089 |
| | emax1 | 0.334 | 0.175 | 0.067 | 0.168 | 0.070 | 0.158 | 0.068 | 0.079 | 0.052 | 0.074 | 0.079 |
| | emax2 | 0.083 | 0.450 | 0.239 | 0.430 | 0.229 | 0.407 | 0.216 | 0.303 | 0.162 | 0.238 | 0.136 |
| | exponential1 | 0.916 | -0.385 | 0.186 | -0.365 | 0.176 | -0.367 | 0.180 | -0.325 | 0.170 | -0.268 | 0.143 |
| | quadratic1 | 0.216 | 0.357 | 0.165 | 0.314 | 0.149 | 0.260 | 0.124 | 0.135 | 0.072 | 0.078 | 0.065 |
| | logistic1 | 0.713 | -0.217 | 0.084 | -0.211 | 0.084 | -0.208 | 0.085 | -0.180 | 0.080 | -0.168 | 0.091 |
| | exponential2 | 0.828 | -0.316 | 0.137 | -0.303 | 0.132 | -0.309 | 0.136 | -0.343 | 0.185 | -0.261 | 0.143 |
| | quadratic2 | 0.257 | 0.273 | 0.114 | 0.252 | 0.107 | 0.236 | 0.102 | 0.114 | 0.080 | 0.084 | 0.072 |
| | sigEmax | 0.291 | 0.209 | 0.081 | 0.190 | 0.077 | 0.185 | 0.074 | 0.139 | 0.082 | 0.100 | 0.075 |
| | power | 0.360 | 0.153 | 0.060 | 0.149 | 0.063 | 0.137 | 0.062 | 0.096 | 0.076 | 0.045 | 0.081 |
| | logistic2 | 0.601 | -0.109 | 0.048 | -0.104 | 0.049 | -0.112 | 0.051 | -0.143 | 0.071 | -0.117 | 0.081 |
| | betaMod | 0.075 | 0.545 | 0.341 | 0.447 | 0.268 | 0.355 | 0.217 | 0.135 | 0.059 | 0.141 | 0.080 |
| $N = 20$ | linear | 0.600 | -0.049 | 0.035 | -0.052 | 0.036 | -0.059 | 0.039 | -0.077 | 0.055 | -0.082 | 0.067 |
| | emax1 | 0.334 | 0.225 | 0.083 | 0.211 | 0.081 | 0.201 | 0.081 | 0.123 | 0.072 | 0.099 | 0.073 |
| | emax2 | 0.083 | 0.512 | 0.295 | 0.470 | 0.268 | 0.438 | 0.252 | 0.300 | 0.156 | 0.258 | 0.134 |
| | exponential1 | 0.916 | -0.327 | 0.141 | -0.303 | 0.129 | -0.296 | 0.129 | -0.241 | 0.117 | -0.196 | 0.087 |
| | quadratic1 | 0.216 | 0.417 | 0.213 | 0.317 | 0.165 | 0.295 | 0.156 | 0.124 | 0.067 | 0.083 | 0.051 |
| | logistic1 | 0.713 | -0.178 | 0.063 | -0.165 | 0.061 | -0.165 | 0.063 | -0.135 | 0.067 | -0.100 | 0.055 |
| | exponential2 | 0.828 | -0.260 | 0.101 | -0.256 | 0.101 | -0.248 | 0.099 | -0.231 | 0.106 | -0.186 | 0.088 |
| | quadratic2 | 0.257 | 0.322 | 0.139 | 0.292 | 0.132 | 0.271 | 0.122 | 0.165 | 0.082 | 0.102 | 0.061 |

**Table S4 continued from previous page**

| | | | | | | | | | | | | |
|---|---|---|---|---|---|---|---|---|---|---|---|---|
| | sigEmax | 0.291 | 0.250 | 0.095 | 0.227 | 0.086 | 0.219 | 0.088 | 0.142 | 0.071 | 0.109 | 0.063 |
| | power | 0.360 | 0.210 | 0.077 | 0.198 | 0.078 | 0.184 | 0.077 | 0.095 | 0.069 | 0.095 | 0.077 |
| | logistic2 | 0.601 | -0.069 | 0.036 | -0.066 | 0.038 | -0.069 | 0.038 | -0.101 | 0.055 | -0.075 | 0.059 |
| | betaMod | 0.075 | 0.586 | 0.392 | 0.332 | 0.191 | 0.267 | 0.154 | 0.117 | 0.044 | 0.136 | 0.055 |
| $N = 30$ | linear | 0.600 | -0.026 | 0.029 | -0.031 | 0.032 | -0.036 | 0.034 | -0.052 | 0.049 | -0.053 | 0.054 |
| | emax1 | 0.334 | 0.254 | 0.095 | 0.227 | 0.088 | 0.220 | 0.089 | 0.145 | 0.078 | 0.118 | 0.070 |
| | emax2 | 0.083 | 0.532 | 0.317 | 0.475 | 0.279 | 0.442 | 0.259 | 0.288 | 0.154 | 0.257 | 0.123 |
| | exponential1 | 0.916 | -0.287 | 0.114 | -0.259 | 0.103 | -0.258 | 0.105 | -0.209 | 0.094 | -0.149 | 0.057 |
| | quadratic1 | 0.216 | 0.391 | 0.200 | 0.280 | 0.152 | 0.266 | 0.145 | 0.108 | 0.057 | 0.096 | 0.047 |
| | logistic1 | 0.713 | -0.156 | 0.052 | -0.141 | 0.052 | -0.136 | 0.051 | -0.104 | 0.051 | -0.086 | 0.043 |
| | exponential2 | 0.828 | -0.234 | 0.084 | -0.223 | 0.082 | -0.225 | 0.084 | -0.195 | 0.088 | -0.154 | 0.066 |
| | quadratic2 | 0.257 | 0.346 | 0.154 | 0.295 | 0.136 | 0.281 | 0.130 | 0.128 | 0.065 | 0.121 | 0.063 |
| | sigEmax | 0.291 | 0.265 | 0.100 | 0.235 | 0.090 | 0.230 | 0.091 | 0.154 | 0.065 | 0.117 | 0.057 |
| | power | 0.360 | 0.235 | 0.086 | 0.216 | 0.082 | 0.201 | 0.079 | 0.126 | 0.071 | 0.096 | 0.066 |
| | logistic2 | 0.601 | -0.045 | 0.030 | -0.047 | 0.032 | -0.046 | 0.033 | -0.056 | 0.044 | -0.045 | 0.046 |
| | betaMod | 0.075 | 0.500 | 0.318 | 0.210 | 0.100 | 0.189 | 0.107 | 0.093 | 0.032 | 0.131 | 0.044 |
| $N = 50$ | linear | 0.600 | -0.006 | 0.024 | -0.005 | 0.027 | -0.004 | 0.028 | -0.031 | 0.039 | -0.031 | 0.042 |
| | emax1 | 0.334 | 0.265 | 0.097 | 0.233 | 0.089 | 0.233 | 0.092 | 0.160 | 0.072 | 0.131 | 0.063 |
| | emax2 | 0.083 | 0.536 | 0.326 | 0.448 | 0.265 | 0.429 | 0.255 | 0.255 | 0.127 | 0.268 | 0.123 |
| | exponential1 | 0.916 | -0.243 | 0.085 | -0.206 | 0.073 | -0.206 | 0.074 | -0.152 | 0.061 | -0.114 | 0.035 |
| | quadratic1 | 0.216 | 0.317 | 0.159 | 0.192 | 0.099 | 0.166 | 0.088 | 0.095 | 0.041 | 0.111 | 0.044 |
| | logistic1 | 0.713 | -0.133 | 0.042 | -0.110 | 0.039 | -0.108 | 0.040 | -0.064 | 0.034 | -0.056 | 0.028 |

**Table S4 continued from previous page**

| | | | | | | | | | | | | |
|---|---|---|---|---|---|---|---|---|---|---|---|---|
| | exponential2 | 0.828 | -0.199 | 0.066 | -0.185 | 0.062 | -0.182 | 0.063 | -0.155 | 0.061 | -0.120 | 0.046 |
| | quadratic2 | 0.257 | 0.338 | 0.154 | 0.266 | 0.124 | 0.254 | 0.122 | 0.140 | 0.064 | 0.124 | 0.052 |
| | sigEmax | 0.291 | 0.264 | 0.097 | 0.232 | 0.089 | 0.230 | 0.088 | 0.152 | 0.060 | 0.131 | 0.052 |
| | power | 0.360 | 0.253 | 0.091 | 0.231 | 0.089 | 0.222 | 0.086 | 0.130 | 0.073 | 0.120 | 0.064 |
| | logistic2 | 0.601 | -0.030 | 0.024 | -0.025 | 0.026 | -0.029 | 0.026 | -0.029 | 0.036 | -0.032 | 0.034 |
| | betaMod | 0.075 | 0.312 | 0.139 | 0.146 | 0.046 | 0.099 | 0.028 | 0.076 | 0.017 | 0.151 | 0.042 |
| $N = 60$ | linear | 0.600 | -0.046 | 0.035 | -0.048 | 0.036 | -0.054 | 0.038 | -0.020 | 0.036 | -0.018 | 0.037 |
| | emax1 | 0.334 | 0.227 | 0.084 | 0.205 | 0.078 | 0.198 | 0.080 | 0.149 | 0.070 | 0.136 | 0.060 |
| | emax2 | 0.083 | 0.506 | 0.290 | 0.464 | 0.262 | 0.424 | 0.240 | 0.233 | 0.116 | 0.273 | 0.119 |
| | exponential1 | 0.916 | -0.328 | 0.141 | -0.307 | 0.132 | -0.299 | 0.131 | -0.136 | 0.050 | -0.104 | 0.029 |
| | quadratic1 | 0.216 | 0.401 | 0.200 | 0.315 | 0.159 | 0.283 | 0.152 | 0.089 | 0.037 | 0.112 | 0.042 |
| | logistic1 | 0.713 | -0.180 | 0.064 | -0.165 | 0.062 | -0.161 | 0.062 | -0.059 | 0.030 | -0.049 | 0.025 |
| | exponential2 | 0.828 | -0.259 | 0.100 | -0.250 | 0.098 | -0.253 | 0.103 | -0.139 | 0.053 | -0.110 | 0.040 |
| | quadratic2 | 0.257 | 0.328 | 0.142 | 0.288 | 0.127 | 0.269 | 0.122 | 0.130 | 0.060 | 0.135 | 0.054 |
| | sigEmax | 0.291 | 0.249 | 0.094 | 0.226 | 0.087 | 0.217 | 0.086 | 0.147 | 0.056 | 0.133 | 0.048 |
| | power | 0.360 | 0.212 | 0.078 | 0.193 | 0.073 | 0.179 | 0.073 | 0.132 | 0.071 | 0.137 | 0.064 |
| | logistic2 | 0.601 | -0.061 | 0.036 | -0.064 | 0.037 | -0.069 | 0.039 | -0.025 | 0.029 | -0.023 | 0.031 |
| | betaMod | 0.075 | 0.603 | 0.406 | 0.342 | 0.207 | 0.221 | 0.124 | 0.072 | 0.014 | 0.154 | 0.042 |

Table S4: Bias and Mean Square Error (MSE) of minimum effective doses (MED's) estimated through LiMAP-curvature, smoothing spline and MCP-Mod across different true underlying dose-response models and sample sizes with the clinical relevance threshold 0.3.

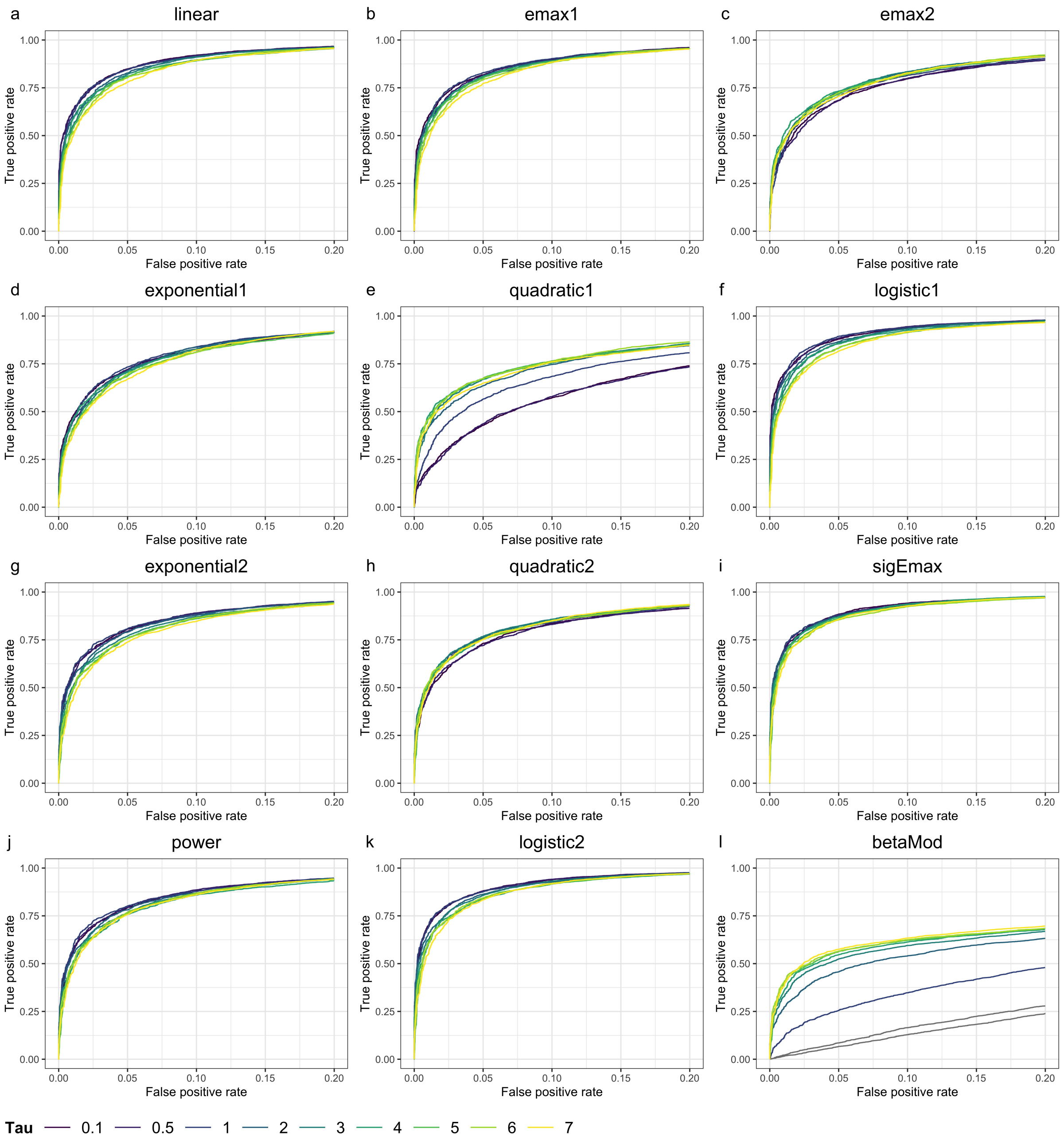


Figure S12: ROC curves of LiMAP-curvature with varying values of $\tau$ across different true underlying dose-response models with the sample size of 40 patients per arm.

| Model | True MED | LiMAP-curvature | | | | | | | | | | | | | | | | | |
|---|---|---|---|---|---|---|---|---|---|---|---|---|---|---|---|---|---|---|---|
| | | $\tau = 0.1$ | | $\tau = 0.5$ | | $\tau = 1$ | | $\tau = 2$ | | $\tau = 3$ | | $\tau = 4$ | | $\tau = 5$ | | $\tau = 6$ | | $\tau = 7$ | |
| | | Bias | MSE | Bias | MSE | Bias | MSE | Bias | MSE | Bias | MSE | Bias | MSE | Bias | MSE | Bias | MSE | Bias | MSE |
| linear | 0.600 | -0.016 | 0.026 | -0.012 | 0.026 | -0.013 | 0.026 | -0.013 | 0.028 | -0.015 | 0.029 | -0.017 | 0.029 | -0.016 | 0.031 | -0.019 | 0.031 | -0.018 | 0.030 |
| emax1 | 0.334 | 0.265 | 0.096 | 0.264 | 0.097 | 0.258 | 0.095 | 0.244 | 0.091 | 0.236 | 0.090 | 0.233 | 0.091 | 0.233 | 0.092 | 0.230 | 0.092 | 0.228 | 0.091 |
| emax2 | 0.083 | 0.571 | 0.353 | 0.564 | 0.346 | 0.537 | 0.323 | 0.493 | 0.295 | 0.463 | 0.273 | 0.450 | 0.267 | 0.439 | 0.261 | 0.435 | 0.258 | 0.435 | 0.258 |
| exponential1 | 0.916 | -0.281 | 0.107 | -0.279 | 0.105 | -0.261 | 0.097 | -0.243 | 0.091 | -0.238 | 0.089 | -0.230 | 0.087 | -0.229 | 0.088 | -0.227 | 0.087 | -0.230 | 0.090 |
| quadratic1 | 0.216 | 0.498 | 0.273 | 0.484 | 0.264 | 0.364 | 0.190 | 0.263 | 0.138 | 0.234 | 0.123 | 0.221 | 0.119 | 0.218 | 0.119 | 0.208 | 0.116 | 0.197 | 0.111 |
| logistic1 | 0.713 | -0.148 | 0.048 | -0.149 | 0.048 | -0.141 | 0.047 | -0.137 | 0.047 | -0.127 | 0.046 | -0.120 | 0.044 | -0.120 | 0.044 | -0.119 | 0.044 | -0.119 | 0.044 |
| exponential2 | 0.828 | -0.220 | 0.076 | -0.221 | 0.076 | -0.214 | 0.074 | -0.207 | 0.073 | -0.203 | 0.072 | -0.200 | 0.073 | -0.199 | 0.071 | -0.201 | 0.073 | -0.201 | 0.073 |
| quadratic2 | 0.257 | 0.379 | 0.171 | 0.378 | 0.171 | 0.346 | 0.155 | 0.300 | 0.138 | 0.283 | 0.131 | 0.270 | 0.127 | 0.269 | 0.128 | 0.268 | 0.129 | 0.264 | 0.127 |
| sigEmax | 0.291 | 0.278 | 0.103 | 0.278 | 0.103 | 0.267 | 0.100 | 0.248 | 0.095 | 0.237 | 0.092 | 0.233 | 0.091 | 0.230 | 0.089 | 0.232 | 0.091 | 0.228 | 0.090 |
| power | 0.360 | 0.251 | 0.090 | 0.249 | 0.089 | 0.244 | 0.088 | 0.232 | 0.086 | 0.228 | 0.087 | 0.223 | 0.086 | 0.223 | 0.087 | 0.218 | 0.086 | 0.212 | 0.084 |
| logistic2 | 0.601 | -0.037 | 0.027 | -0.037 | 0.027 | -0.037 | 0.027 | -0.032 | 0.028 | -0.035 | 0.029 | -0.032 | 0.029 | -0.033 | 0.029 | -0.036 | 0.029 | -0.032 | 0.029 |
| betaMod | 0.075 | 0.721 | 0.540 | 0.661 | 0.478 | 0.375 | 0.201 | 0.233 | 0.103 | 0.183 | 0.079 | 0.156 | 0.066 | 0.142 | 0.063 | 0.139 | 0.062 | 0.132 | 0.061 |

Table S5: Bias and Mean Square Error (MSE) of minimum effective doses (MED's) estimated through LiMAP-curvature with varying values of $\tau$ across different true underlying dose-response models with the sample size of 40 patients per arm with the clinical relevance threshold 0.3.
